# Person, place, or preference? Combining subjective and objective decision agency with choice preferences predicts behavior during major shifts in human mobility

**DOI:** 10.1101/2025.09.08.25335351

**Authors:** Qing Yao, Sarah Ashcroft-Jones, Sen Pei, Kai Ruggeri

## Abstract

Human mobility plays a crucial role in shaping outcomes such as disease transmission dynamics, economic trends, and access to critical services. In this study, we explored the interplay between decision preferences, public health policy, and mobility patterns during a recent, near-universal shift in human behavior: the COVID-19 pandemic in New York City. We surveyed N = 1049 residents capturing three key decision-making preferences: temporal discounting, loss aversion, and agency. In tandem, population mobility in 2020 was measured for six different “place” categories (grocery stores/pharmacies, general retail, arts/entertainment, restaurants/bars, education, healthcare) using mobile phone-derived foot-traffic data.

The results indicate that decision agency and choice preferences were significantly correlated with zip code-level per capita visits to different places, accounting for disease spread, policy stringency, and socioeconomic variables. We further assessed the predictive power of the scores via out-of-sample predictions using Random Forest models. The results underscore the importance of capturing these behavioral mechanisms in public health intervention strategies and policy.

## Introduction

Human mobility (Barbosa et al., 2018), a fundamental aspect of human behavior, can undergo profound shifts in response to major disruptions such as technological advancements, environmental crises (W. Li et al., 2022), and public health emergencies (Bajardi et al., 2011). Such events can trigger sweeping changes across facets of human activity, including consumer behavior, work habits, and education, ultimately leading to significant alterations in mobility patterns (Grashuis et al., 2020; Liu et al., 2020; Rajput & Mostafavi, 2023; “When the World Stops Moving,” 2020; World Bank, 2020). How these changes occur depends on the interplay between the environment, behavior, policy, and the event itself. One argument is, therefore, that understanding the mechanisms behind these changes requires examination of established decision patterns and cognitive biases. For example, cognitive biases are thought to play a role in voluntary non-migration from stressful environments (Czaika & Reinprecht, 2022), and appear to also influence more everyday mobility choices such as using private over public transportation (Innocenti et al., 2013).

Among recent global disruptions, the COVID-19 pandemic stands out as being particularly impactful on human behavior and mobility. Indeed, a substantial body of research has since analyzed how public health intervention policies during the pandemic shaped individual- and population-level mobility (Byrne et al., 2021; Kraemer et al., 2020; Nouvellet et al., 2021; Pedrosa et al., 2020; Pullano et al., 2020). Empirical studies further demonstrated the interplay between this change in mobility and disease spread, with evidence that mobility reductions during the early phase of the pandemic correlated with lower SARS-CoV-2 transmission rates (Badr et al., 2020; Kissler et al., 2020; Nouvellet et al., 2021). Studies also underscored the importance of integrating choice preferences and behavioral dynamics into predictive models for infectious diseases, emphasizing that understanding the behavioral mechanisms influencing mobility changes is essential for improving intervention strategies (Bedson et al., 2021; Hajlasz & Pei, 2024). Indeed, adherence to and compliance with mobility relevant policy such as shelter-in-place measures, adopted by many countries globally, is known to be explained, at least in part, by psychological traits including agreeableness (Chan et al., 2021). However, it is less clear how other psychological constructs such as cognitive biases may inform understanding of shifts in human mobility during times of crisis. Furthermore, it remains relatively unexplored how the influence of these biases may be further shaped by the environment of the decision maker. Indeed, the related interplay between green spaces in urban settings and the impact of COVID-19 on population (Maury-Mora et al., 2022; Ugolini et al., 2021), for example, indicates how the structure of an environment may help or hinder adherence to public health measures such as lockdowns and shelter-in-place orders.

Human decision makers often appear irrational when making decision under uncertainty, with suboptimal choices arising as a result of biases in cognition (Tversky & Kahneman, 1974). These biases are impactful, with several established cognitive biases, such as temporal discounting (Ruggeri et al., 2022) and risk aversion (Rudisill, 2013), being predictive of broad human behaviors across domains. Importantly, these bias measures are indicative of broader choice preferences, tendencies towards choice types in the decision landscape. This kind of choice preference is evident in many contexts, including general mobility choices (Adam, 2024) and that of the COVID-19 pandemic itself. For example, in one multi-country study, individuals with higher levels of temporal discounting (i.e., the tendency to place more weight on immediate factors than on future ones) were less likely to get vaccinated (Halilova et al., 2022). By contrast, framing vaccination communications with reference to potential losses due to non-vaccination leveraged the loss aversion choice preference (e.g. losses loom larger than gains) to improve parental acceptance of COIVD-19 vaccination (Wang et al., 2022). In another study examining mask-wearing behaviors, loss framing increased support for masking, moderated by political ideology (Steffen & Cheng, 2021). This effect depended on who the message appealed to: loss-framing was most persuasive when distant social entities were evoked – such as family and friends, rather than the self (Jiang & Dodoo, 2021). Another choice preference is risk sensitivity and research noted that higher infection risk perception may be associated with increased adoption of personal hygiene behavior (Schumpe et al., 2022). Speaking directly to changes in mobility, risk aversion tendencies were found to predict reduction in mobility for six location-specific mobility measures for 132 countries based on geocoded Global Preferences Survey (Chan et al., 2020). These works evidence the importance of considering choice preferences, as captured in cognitive bias measures, in understanding mobility trends and behaviors in times of uncertainty and global upheaval.

Another but often under acknowledged aspect of decision-making, which constrains these choice preferences, is decision agency. Broadly speaking agency speaks to an individual’s capacity to take actions based on their own free will, desires, and intentions (Banerjee et al., 2024). In terms of decision making, agency can be described in relation to an individual’s choice set, with a larger set of options relating to higher agency (Sen, 1988). This description makes the environment of the decision maker explicitly relevant to their behaviour. Indeed, even if an individual has free will, if their choice set is limited, their choices are constrained to those options which are possible. This is highly relevant to choice preferences; an option must be a possible for a choice preference to be enacted. This interplay is one that this paper seeks to acknowledge explicitly in the context of the socioeconomic landscape of New York City.

Therefore, despite a large body of extant literature, several fundamental questions about shifts in population mobility during major shifts in human behavior remain open: What roles do context specific decision agency and choice preferences play in shaping decision-making in relation to different mobility-relevant activities (e.g., work, dining, and shopping)? How may these behavioral patterns influence human mobility during periods of major upheaval such as the pandemic? In addition to these questions, the concept of agency (Banerjee et al., 2024; Gneezy et al., 2020) has not been systematically measured in direct conjunction with mobility changes during the COVID-19 pandemic.

In this study, we aim to address these questions by examining the relationship between individual decision-making preferences and human mobility in New York City in 2020. We conducted an extensive survey of N = 2,019 long-term New York City residents to quantify individual choice preferences, including constructs such as temporal discounting and loss aversion. Additionally, we designed and validated a novel measure of decision agency as related to relevant behaviors, including public transportation usage, during lockdown in 2020. This new agency score captures both the availability and practicality/accessibility of these services in 2020. By exploring both these objective and subjective aspects of agency, it offers a contextualized understanding of the key environmental factors affecting the decision agency of New York City residents. To consolidate our insights, behavioral measures were adapted into scores, which were aggregated from the 1049 participants across 17 specifically targeted zip codes that had sufficient representation. These zip codes spanned all five boroughs to allow for an examination of the differences in contextual factors as relevant to specific neighborhoods and zip codes.

To capture travel patterns of essential and non-essential daily activities, as well as varied behavioral responses to pandemic conditions as relevant to individuals’ decision-making agency and biases, we measured population mobility in 2020 across six “place” categories: essential retail (grocery stores and pharmacies), general retail, arts and entertainment venues, restaurants and bars, educational institutions, and healthcare facilities. Mobility data was obtained from a mobile phone-derived foot-traffic dataset provided by Advan Research Corporation (*Advan*, n.d.), which aggregates anonymized location data to deliver insights into physical movement patterns.

We found that, after controlling for COVID-19 policy stringency and socioeconomic factors, temporal discounting and loss aversion scores were significantly positively correlated with per 100 capita visits to the six place categories, while significant negative correlations were found between agency scores and mobility in the four “essential needs” travel destinations. Agency did not relate to mobility as relevant to the two non-essential place categories – restaurants and arts/entertainment venues, evidencing the discernment of this measure. Our findings suggest that both choice preferences and decision agency can help to explain human mobility in response to a major shift in human behavior, such as during the pandemic. This underscores the importance of considering behavioral mechanisms as well as contextualized agency in designing intervention policies (Bavel et al., 2020; Bryan et al., 2021).

## Results

### Human mobility at individual and population level

While this study is not specifically aimed at understanding the COVID-19 pandemic, or pandemics more generally, 2020 presents a valuable example of a period of major shift in human mobility during a crisis. In particular, New York City was highly impacted early in the pandemic, in terms of both transmission rates and economic, employment, educational, and social impacts. Thus, this study focuses on human mobility in New York City during this time as a valid period of major mobility upheaval. This section presents an overview of that shift in mobility, and the gradual reversion – but not full return – to similar patterns as prior to March 2020.

In this study, population mobility was calculated using foot traffic data provided by Advan (Safegraph, 2023). This dataset recorded visits and the number of visitors from estimated home locations (at the census block group level) for millions of places of interest (POIs). Using the North American Industry Classification System (NAICS) codes (Bureau, 2023), we selected POIs in six location categories: i. grocery stores and pharmacies, ii. general retail, iii. arts and entertainment venues, iv. restaurants and bars, v. educational settings, and vi. healthcare facilities from this dataset (Table S1). Our analysis focused on these categories because variation in mobility in these POIs reflects the changing intensity of common human activities during the pandemic. To capture the temporal variation of human mobility in 2020, we computed the weekly sum of visits to each POI place category among residents in each zip code in New York City.

The time series of the number of visits to the six POI categories is depicted in Figure 1a. Overall, the total number of visits varies with the different place categories across the study period. In March 2020, there was a sharp decline across all categories, corresponding to the implementation of the lockdown protocols, with ‘educational settings’ showing the most significant decline. A partial recovery followed during the summer reopening phase for five categories. Additionally, mobility patterns also varied significantly between zip codes. The geographical distribution of percentage changes in mobility during the period from March 2020 to February 2021 is shown in Figure 1b-g. Descriptively, the extent of changes and the specific zip codes affected varied across different categories. The largest overall decrease in visits was observed in educational settings, whereas healthcare facilities exhibited more zip codes with smaller changes, highlighting the spatial variability in mobility patterns during the pandemic even within one large urban setting. See *Materials and Methods* for more information about the mobility data processing. Time-varying mobility patterns on maps for different categories and participants’ changes in indoor activities can also be found in the *Supplementary Information* (SI).

**FIGURE 1:**
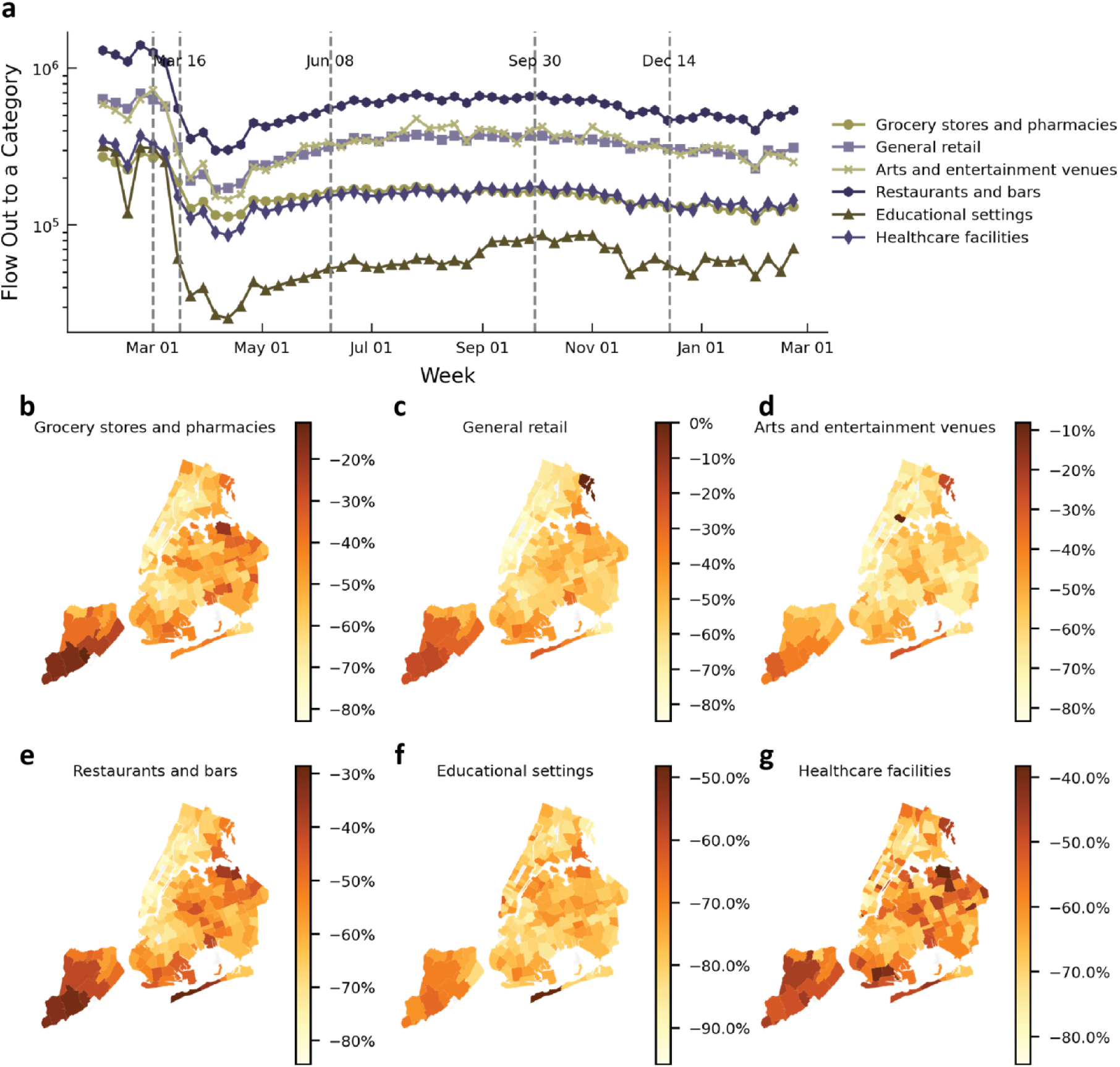
Human mobility patterns during the pandemic. *Note.* Panel **a** shows the temporal trends in weekly number of visits for each category, the dashed vertical lines indicate the major policy changes, for instance, June 8 2020, marked the start of New York City’s reopening. Panels **b–g** demonstrate the geographical distribution of percentages of visit changes from the February 24^th^ 2020 to the February 28^th^ 2021. Note that the values of the visit change scales vary across panels with some of the largest but least varied reductions in mobility occurring in “Educational settings” (panel f), for example.

### Quantifying choice preferences and decision agency

To ensure reliability of participant demographics, we used a paid data collection agency to conduct anonymous online surveys measuring the choice preferences and decision agency of long-term New York City residents. The data collection window was October - December 2023 and was carried out on the Qualtrics platform. A total of 2019 participants from across the five boroughs of New York City responded. After filtering out invalid responses, we retained 1344 valid samples for individual-level analysis, covering 136 zip codes. Detailed survey information, particularly on establishing a subset of focused zip codes, is provided in the Data collection section of the *Materials and Methods*.

The survey was designed to assess residents’ behavioral patterns, including *temporal discounting*, *loss aversion*, and *agency*, as well as personal demographic and socioeconomic characteristics. Temporal discounting was evaluated through a dynamic series of financial choice scenarios, while loss aversion was assessed via preferences for different monetary amounts under various levels of uncertainty (Ruggeri et al., 2022, 2023).

Agency was measured using a newly developed instrument tailored to the pandemic context. Specifically, participants evaluated nine services they would potentially have used during March 2020 to February 2021, such as ride-sharing with Uber or engaging in remote work. Uniquely, each service was assessed first as whether it was objectively available and then second, whether it was subjectively accessible. This produced a total of 18 individual agency measures as participants first indicated whether a particular service existed in their context (e.g., subway, food delivery services) and then rated how realistic it was for them to use (e.g., affordable, safe). This aimed to capture the interaction between the behavior of the individual and their specific urban environment within New York City.

We established both measures intentionally: while there is a correlation between availability and accessibility, the former does not guarantee the latter. For example, Uber might exist in a neighborhood but may not be feasible as a common transport method due to cost. By comparison, subway lines may not readily connect to certain areas, but they would be financially accessible to many if established. Thus, both aspects of agency are independently informative and can be compared against the objective mobility data. Indeed, knowing what exists within proximity to an individual indicates whether a specific service is in the locale or what might be sparsely available to residents. Knowing the subjective perception of accessibility indicates whether it would be used if accessibility was improved. (A third level, utility based on conditional availability, was considered but determined not to be relevant in this context. It would inform likelihood of use if a service were introduced). This new agency measure was validated by assessing its internal consistency, reliability, and by confirming an overarching one-factor model using factor analysis, as detailed in the Survey data section of the *Materials and Method*s section.

Based on these measures, in our survey, a higher score for temporal discounting represents a higher tendency to prefer immediate rewards over delayed gains, and a higher score for loss aversion reflects greater psychological reluctance to accept losses compared with the same amount of gain. Finally, a higher score for agency indicates better access (objectively and subjectively) to necessary resources, facilities, and services during lockdown within the context of New York City.

### Objective and subjective decision agency

The newly constructed agency score focuses on two key dimensions: objective availability and subjective accessibility of decision agency during lockdowns in the specific environmental context of New York. These two facets capture both objective (e.g., "Can I afford or use this as I want?") and subjective (e.g., "Is this accessible to me?") aspects of the possibilities that agents can exercise. The differences between these dimensions reflect the constraints on individuals’ ability to act freely. Figure 2 visualizes these self-reported differences for nine services during the lockdown period in 2020.

**FIGURE 2:**
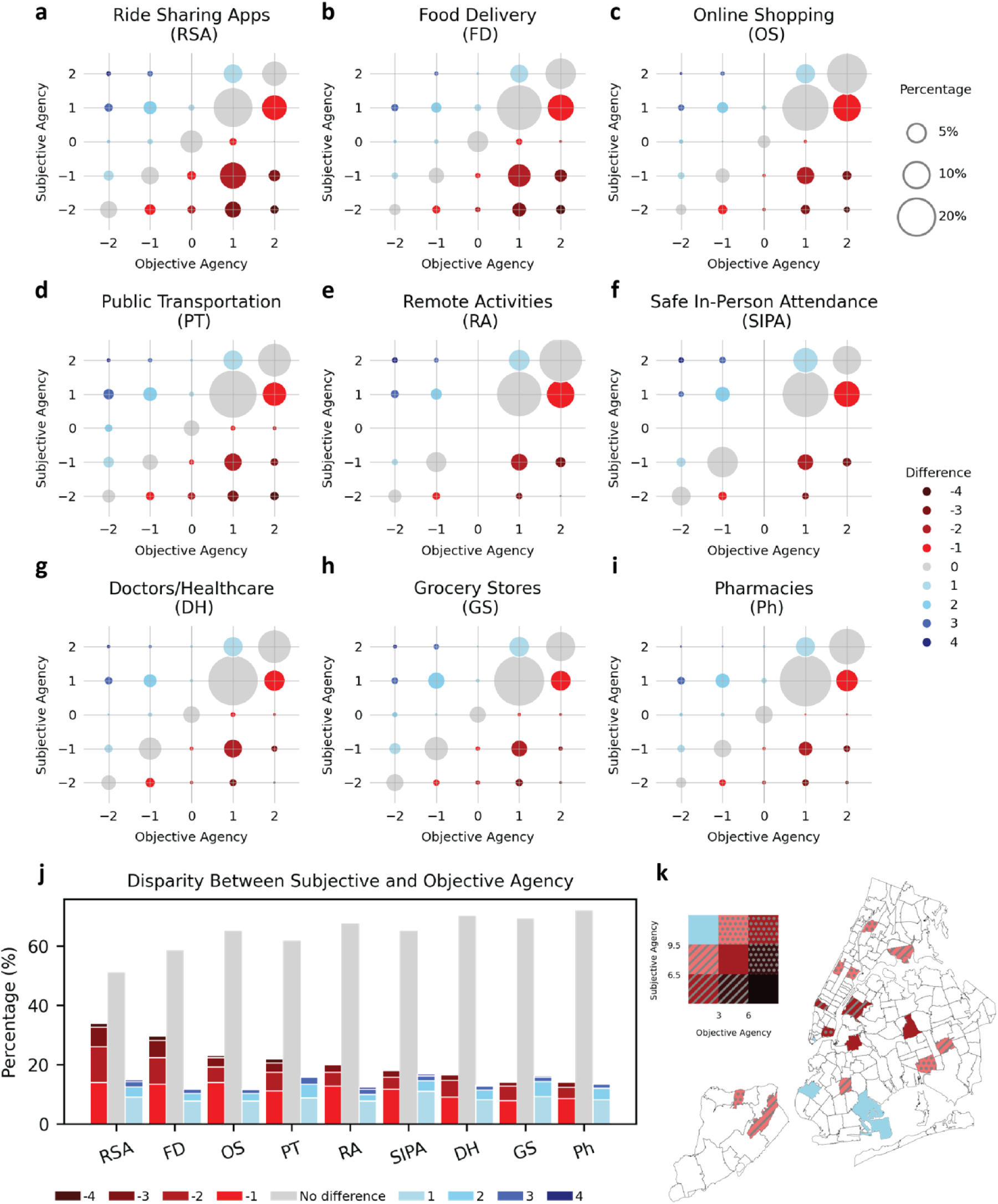
Agency rating across services during lockdowns in NYC. *Note***. a – i**. are bubble plots demonstrating the relationships between objective and subjective agency across different services. The x-axis represents the objective agency while the y-axis represents subjective agency. Both range from -2 to 2. The sizes of the bubble stand for the percentages of participants who chose a certain combination of objective and subjective agency.

As shown in Figure 2, differences between perceived objective and subjective agency were prevalent across all services. Ride-sharing applications, food delivery, online shopping services and public transportation exhibited the highest percentages of participants who rated their objective agency and subjective agency differently (see panels a-f in Figure 2). In contrast, essential services such as grocery stores, pharmacies, and doctors or other healthcare had fewer differences between objective and subjective agency (see panels g-i in Figure 2). Importantly, these differences were not symmetrical — cases where services were considered objectively available but subjectively less accessible (objective > subjective, colored in red in Figure 2) were more common.

The three colors represent three groups: shades of red (negative difference: Objective agency > Subjective agency) indicate that service is available (objective) but not realistic to use (subjective); grey (no difference: Objective agency ∼ Subjective agency) reflects those participants who perceived the service to be equally subjectively available and objectively accessible; shades of blue (positive differences: Objective agency < Subjective agency) indicate a service is subjectively accessible but objectively less available. The differences are represented by nine distinct values ranging from -4 to +4 (see Survey data section in *Materials and methods* for details), with darker colors indicating larger differences. **j.** Bar plot showing the distribution of disparities between the participants’ perceived subjective agency and objective agency of each service. **k.** shows the bivariate color scheme of aggregated objective and subjective agency in the ZIP codes where at least 15 participants provided responses. The colors represent differences in objective and subjective agency levels, following the same rules as the previous subpanels. And the patterns (e.g., stripes and dots) are used to differentiate between similar levels of difference but varying overall levels of agency. Higher overall agencies are presented by the dots while the lower agencies are illustrated with stripes.

The difference between subjectivity and objectivity in agency scores showed substantial spatial variations among New York City zip codes (Figure 2k). For instance, in wealthier neighborhoods like the Upper West Side (10025), decision agencies were high and more balanced for most services, suggesting fewer constraints on the ability to access these services during the lockdown. This directly speaks to the interplay between behavior and context – what is possible sets the bounds on what behaviors can be observed. The differences between subjectivity and objectivity in agency for each service across New York City neighborhoods can be found in the SI.

### Behavioral scores at individual level

After data processing and scoring, the three behavioral scores—temporal discounting, loss aversion, and agency— were found to vary with distinct minimum and maximum values, as detailed in Table S1 of the SI. For comparative analysis, we scaled the range of possible score values to be [0, 100].

To test the consistency of our measured scores against previous studies, we performed a linear regression to examine how the three behavioral scores vary with participants’ financial status as scored through a survey item assessing their financial situation in 2020. The question asked: “Which best describes your financial situation during March-December 2020?” with responses ranging from “unable to pay my bills (scored 1)” to “my bills were paid off each month, and I have enough to save, invest, or spend freely (scored 6)”. For comparability with other individual-level information that had varying numbers of response options, we scaled the range of possible answers to [0, 100]. See Table 1 and Figure 3 for the regression results. As indicated in Figs. 3a and 3c, temporal discounting and agency scores exhibited a negative (*β* = −0.063, *p* < 0.001) and positive (*β* = 0.015, *p* < 0.001) correlation with individuals’ financial status, respectively. These findings align with the general understanding that individuals with better financial conditions tend to have lower temporal discounting (Green et al., 1996) and have a stronger capacity to act on a greater number of available choices (i.e., greater agency) (Deci & Ryan, 2000). However, loss aversion scores did not show significant correlation (*p* = 0.625) between financial statuses among the participants (see Figure1b). A similar result examining loss aversion and financial status can be found in a recent large-scale multi-county study (Ruggeri et al., 2023). Additional details on other individual measurements and their correlations with the scores are available in the SI.

**FIGURE 3:**
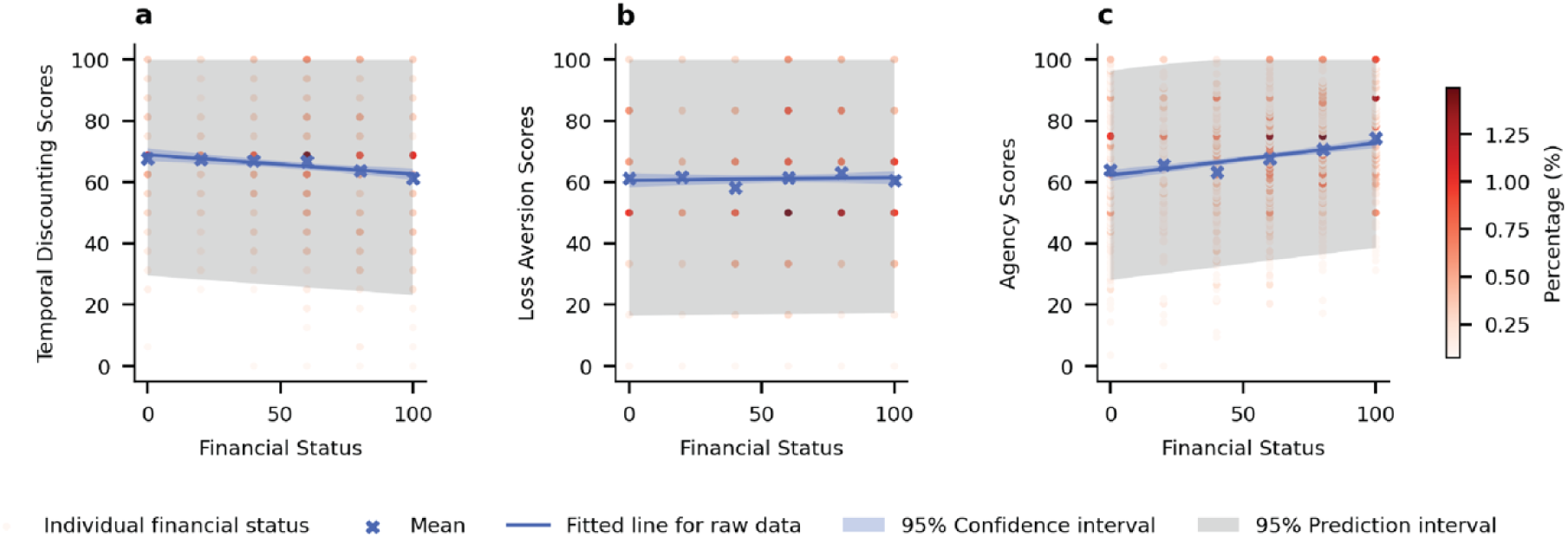
Regression analysis of financial status on behavioral scores. *Note.* Panels **a – c** display scatter plots illustrating the relationship between financial status and individual scores (temporal discounting, loss aversion, and agency). Red dots represent individuals and their corresponding scores; darker red dots indicate a higher density of participants at specific financial statuses and behavioral scores (here N = 1,341 the maximum number of valid samples available to evaluate the overall study is N = 1344, however, N = 3 of those responses did not contain complete financial status reports so were removed from this figure). Blue crosses indicate the mean score values for certain financial status. The blue lines show the ordinary least squares (OLS) regression fits, with shaded blue and grey areas representing the 95% confidence intervals and 95% prediction intervals, respectively.

**Table 1.** OLS Regression Results.

|  | *Temporal discounting* | *Loss aversion* | *Agency* |
| --- | --- | --- | --- |
| *Financial Status-* | -0.063 *** | 0.0092 | 0.11 *** |
| *Coefficient, b* | (0.017) | (0.019) | (0.015) |
| *Intercept* | 68.94 *** | 60.58 *** | 62.23 *** |
|  | (1.09) | (1.21) | (0.94) |
| *R-squared* | 0.01 | 0 | 0.037 |
| *F-statistic* | 13.98 | 0.24 | 51.62 |
| *Prob (F-statistic)* | 0.00019 | 0.63 | 1.11 × 10<sup>-12</sup> |

*Note.* Each column represents an OLS regression model analyzing the relationship between financial status and behavioral score (temporal discounting, loss aversion, or agency).
Standard errors are reported in parentheses. \*\*\* indicates $p$ -values < 0.001.

### Behavioral scores at population level

We next aimed to evaluate whether choice preferences and decision agency predicted the population-level mobility patterns to the POIs identified. To do this, we computed the three behavioral scores at the geographical scale of modified zip-code covered areas (MODZCTA, here after referred to as zip codes, see *Materials and Methods* for details). To ensure that the survey results can reasonably represent the overall decision patterns for a zip code, in the following statistical analysis, we only used the data from zip codes with at least 15 participants. The 17 valid zip codes spanned all five boroughs of New York City, capturing the diverse socioeconomic conditions across New York City neighborhoods and represent a total of N =1049 individual survey responses.

We observed variations in all three behavioral scores across these 17 zip codes, despite a relatively large within-zip-code variation (Figure 4d-f). To avoid potential biased measurement driven by outliers in zip codes with relatively low participant numbers, we applied kernel density estimation to estimate the distributions of behavioral scores in each zip code and used the modes to represent the zip-code level scores.

**FIGURE 4:**
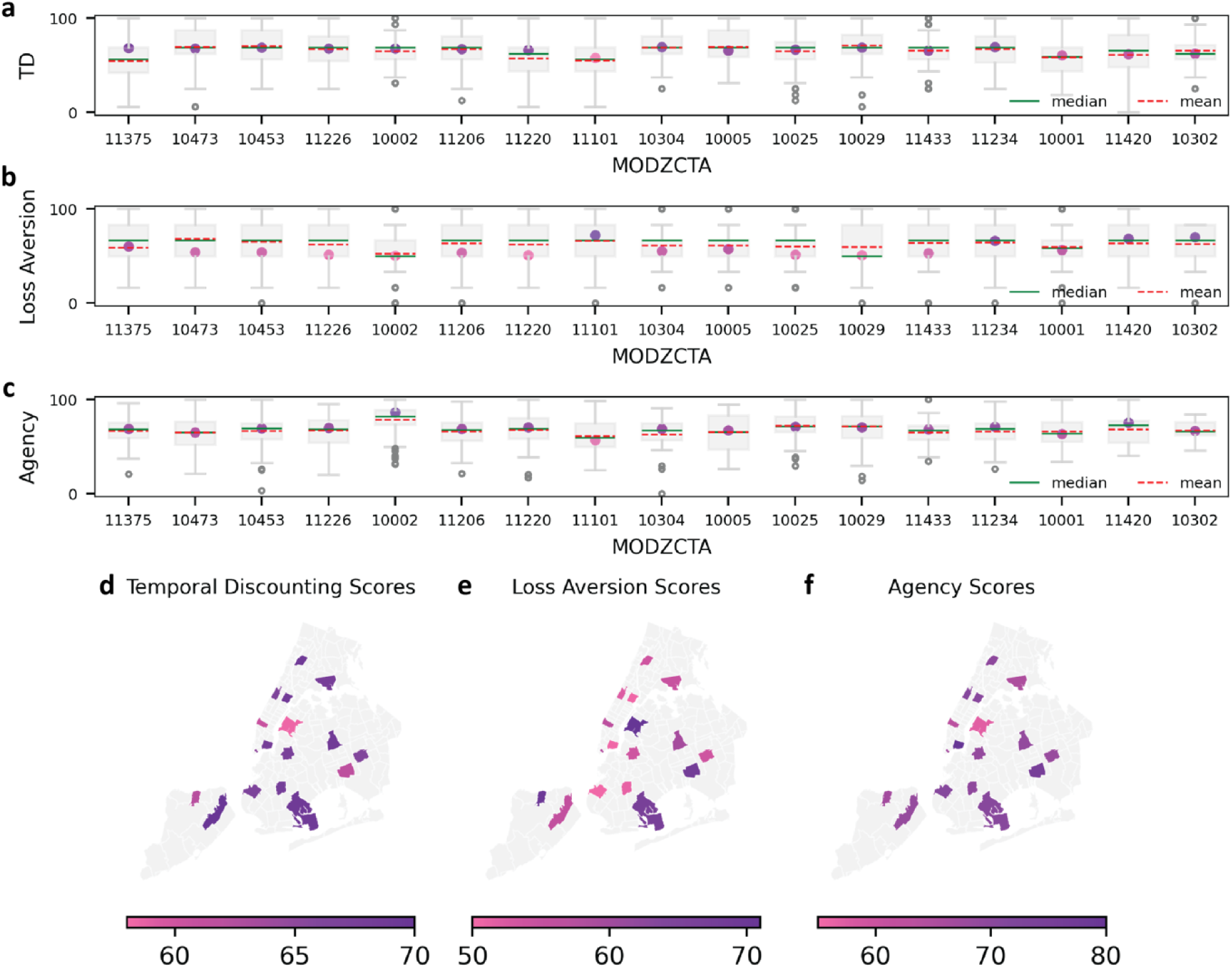
Distributions and variation in behavior scores across zip codes. *Note.* Panels **a** – **c** illustrates the distributions of the scores in the zip codes with at least 15 valid participant responses. The boxes show the median (green solid line), mean (red dashed line) and interquartile range (IQR) of distributions. Whiskers show the range of 1.5 IQR below the first quartile and above the third quartile. The empty circles indicate the data points outside this range. The modes of kernel density estimations (KDE) are shown by the filled dots. Panels **d – f** presents the geographical distribution of modes values of scores for temporal discounting, loss aversion, and agency across New York City zip codes with at least 15 participants. Areas with not enough participants for inclusion in this analysis, or no valid participants, are shown in light grey. A higher temporal discounting score indicates a stronger preference for immediate rewards over delayed gains. Higher loss aversion scores reflect greater reluctance to accept losses compared to equivalent gains. Higher agency scores signify better availability of and access to essential resources, facilities, and services during lockdown.

### Behavioral scores and mobility

To explore the behavioral mechanisms as relevant to the population-level mobility during the COVID-19 pandemic, we then applied a generalized additive model (GAM) to estimate how the behavioral scores predicted weekly visitor numbers per 100 population to the six place categories in the 17 valid zip codes. In the analysis, we focus on the period from March 16^th^, 2020, to February 28^th^, 2021 (50 weeks in total). As the mobility of residents in these activities was likely heavily impacted by socio-economic conditions, we controlled for a range of covariates including percentage of population with no health insurance, percentage of households with no vehicles, median value of household income, and population weighted average ages. In addition, we included the weekly number of COVID-19 cases for each of the five boroughs of New York City, with the case count for each zip code corresponding to the borough in which that zip code is located, to represent the perceived infection risk in the city as relevant to the zip code itself, the Oxford government stringency index was included to capture the strictness of intervention policies (Hale et al., 2020), and the number of weeks since March 16, 2020, was used to measure the potential impact of “pandemic fatigue” (Haktanir et al., 2022). The model specifications are detailed in the Generalized Additive Model subsection of *Materials and Methods*. A comprehensive description of the dataset, full model results and model validation are provided in the SI.

Across all place categories, the temporal discounting (TD) score showed a persistent positive coefficient ( < 0.05) (top row of Figure 5), holding all other variables constant. This implies that the more participants tended on average to favor immediate rewards (high TD scores), the higher the mobility to all place categories. Similarly, the marginal effect of loss aversion score had a positive coefficient as relevant to mobility in all place categories (*p* < 0.05) (middle row of Figure 5). That is, locations with residents who are more sensitive on average to potential losses showed higher mobility during the pandemic.

**FIGURE 5:**
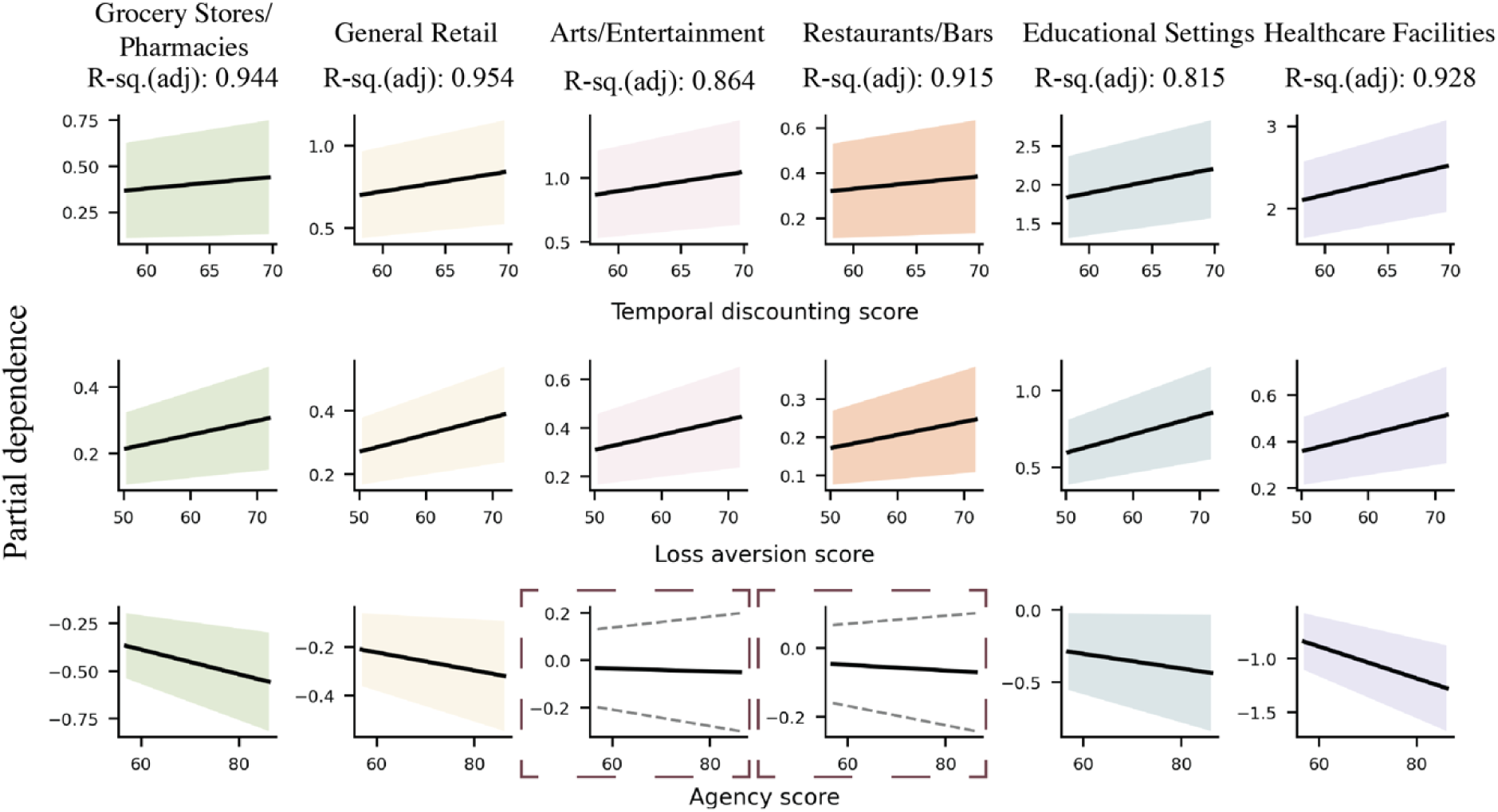
Partial dependence of three behavioral scores for the GAM model. *Note.* This figure presents the GAM results for the three behavioral measures: temporal discounting (row 1), loss aversion (row 2), and agency (row 3). Each column relates to one of the six place categories. The solid lines represent the predicted weekly visits based on the marginal effect of the scores, accounting for the contributions of all other variables in the model. The shaded areas reflect the uncertainty range (95% confidence intervals). The panels in dashed frames show terms that are not statistically significant in the model (*p* > 0.05). Adjusted *R*^2^ shown at the top are the regression results of each full model, see the *Materials and Method*s section for the detailed GAM model structure.

By contrast, the marginal effect of the agency score had a negative coefficient for several place categories (bottom row of Figure 5). In this model, across POIs (except the arts and entertainment venues and restaurants and bars), a higher decision agency predicted lower aggregate mobility. The effects of agency for arts and entertainment as well as restaurants and bars were not significant. This place-based difference in agency may relate to the top-down policies which dictated the closure of “non-essential” venues thereby overriding any influence of individual agency. Speaking to comparisons within the same categories (each column), the TD score generally had a larger regression coefficient indicating a greater change in mobility with change in temporal discounting as compared with either loss aversion or agency.

We constructed Random Forest (RF) regression models to perform out-of-sample predictions of weekly mobility per 100 capita. Since socio-economic variables and behavioral scores are correlated and we concentrated on the behavioral scores for their predictive importance, therefore, the models included only the behavioral scores alongside three temporal predictors: week index, weekly case number, and stringency index. For each category, models were trained using an expanding window approach, forecasting the upcoming week’s mobility across all locations over 19 weeks. To compare the importance of variables in predicting across different models of six categories, we calculated the mean relative SHAP values of the predictions for each variable within each category (see Figure 6). Detailed model specifications and the definition of SHAP importance are provided in the Random Forest Model subsection of the *Materials and Methods*.

**FIGURE 6:**
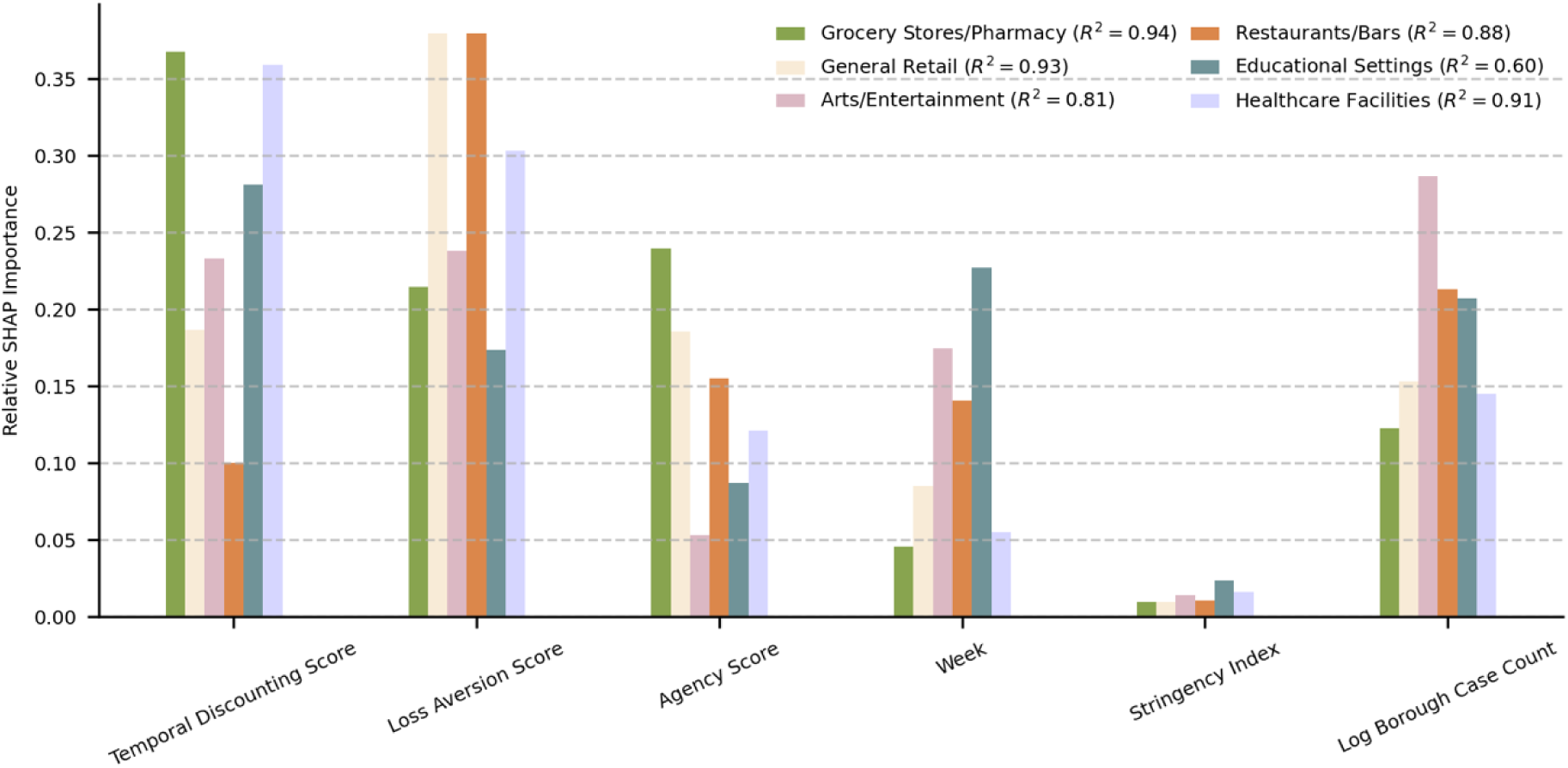
Bar plots of relative SHAP importance of Random Forest models. *Note.* The colors indicating different place categories and the R-squared values for the whole prediction periods are shown in the brackets.

With the scores and three temporal variables, the RF models had a good fit for almost all the categories except for the Educational Settings, indicating the behavioral scores have predictive powers for human mobility. The stringency index showed relatively low importance as it correlated with week index and case counts. The importances pattern varied across the categories: for instance, to predict weekly visits per 100 capita to grocery stores and pharmacies and healthcare facilities, the behavioral scores had higher importance compared with temporal variables, while the predictions of arts and entertainment had high relevance with temporal case counts. The time series plots of prediction and true values in different MODZCTA can be found in the SI. For sensitivity analysis, we performed the RF models with socio-economic data included and no great improvement was observed.

## Discussion

Shifts in human mobility in emergent crises are indicative of responses to ongoing changes in policy, such as evacuation patterns (X. Li et al., 2024), and population-level disaster resilience, such as long term adherence to shelter in place orders. Crises such as natural disasters, extreme weather situations, or outbreaks of infectious disease also inherently challenge the resilience and “insurance value” of urban areas to such mobility shifts (Gómez-Baggethun & Barton, 2013; Venter et al., 2020). Furthermore, these shifts can reveal important dynamics in the interplay between individual choice patterns, environmental factors, decision agency, and mobility behaviors at scale.

To that end, this study linked survey data on contextualized decision agency and choice preferences from established New York City residents with mobile phone-derived foot-traffic data that tracked mobility patterns in New York City during a period of extreme, near-universal shifts in human behavior (the COVID-19 pandemic). Our study quantified choice preferences and decision agency and mapped spatiotemporal mobility trends captured to six different place categories. Our findings revealed that temporal discounting, loss aversion, and agency were each correlated with and were important in predicting visitor numbers in most of the examined place categories, suggesting that agency and choice preferences might help to explain changes in population mobility during a public health emergency. This result also suggests that these scores are widely valuable predictors for decision-making in a wide range of activities during periods of uncertainty.

Understanding how people make mobility decisions during a public crisis is critical for policymaking. Understanding how the context of those choices influences the decision landscape of individuals is equally vital. Our study indicates that residents in different neighborhoods may react to such policies differently, based on characteristics of that subpopulation and the contextualized agency they have in that environment, resulting in different patterns of mobility changes. Such nuance and local contextualization need to be considered when evaluating the potential effects of policy changes. In major crises that result in significant shifts in human mobility, such as airborne pandemics, predicting human mobility change is key. Understanding mobility changes in reaction to infection risk is key to generating reliable mid- and long-term forecasts and to design effective policies and interventions. Our findings suggest possible behavioral factors to be included in future models for predicting mobility during epidemics and highlight that future models should account for both the structural environmental constraints that limit certain behaviors (agency) and the predisposition of individuals toward specific choices (choice preferences). These insights are essential for developing targeted, feasible strategies to improve emergency preparedness and management.

The introduction of our agency score enhances the understanding, relevance, and potential applicability of this study. This measure not only considers what options are objectively available but also what choices are subjectively realistic for individuals, which produced distinct results in noteworthy ways, particularly at the neighborhood level. Doing so has allowed us to link issues of financial, environmental, and social barriers that are vital to understanding decision-making. For instance, options like working from home or avoiding public transit are not accessible to everyone, even during a global pandemic. Our analysis on the differences between subjective agency and objective agency revealed the idea of vulnerable neighborhoods with reduced access to certain services due to resource constraints. In these neighborhoods the stringency index, which represents the strictness of lockdown measures at a given point in time, would not necessarily have been equally easy or reasonable to follow compared to better resourced neighborhoods and subpopulations. Future emergency policy interventions could benefit from more precisely targeting such neighborhoods within urban environments. Indeed, more specific service provisions to ameliorate these socio-economic barriers may also help to improve equity in future health emergencies or similar emergent mobility shifts.

However, this study is not without limitations. The survey on agency and choice preferences was performed two years after the study period of mobility in 2020-2021. The population composition in New York City may have changed in that time, weakening the link between the zip code mobility and behavioral scores. However, by restricting survey participants to a representative sample of residents who had lived in the same zip codes in New York City since 2020, i.e. stable residents of that zip code, we attempted to reduce that source of data noise. Yet, while decision-making preferences are generally considered to be stable cognitive features, the possibility of their change over time remains. Second, for privacy reasons, we were unable to track the individual-level mobility of survey participants. Therefore, we aggregated behavioral scores to the zip code-level and quantified their impact on population-level mobility. Importantly, for infectious disease spread, we argue that aggregated neighborhood-level mobility data can support accurate modeling (Buckee et al., 2020; Zhang et al., 2024). Third, as numbers of participants are low in many zip codes, the analysis is limited to the 17 zip codes which held sufficient sample numbers. Surveys targeting specific zip codes are hard to implement and costly to fund. With greater resources to collect a larger and more distributed sample, we could better quantify zip code-level behavioral scores more accurately and across more New York City neighborhoods. In turn, this would increase understanding of the relationship between local and contextualized barriers to policy adherence (as indicated by the agency measure) and shifts in large-scale mobility trends.

Despite these limitations, our study holds several important findings. Foremost, the creation of our novel contextualized agency measure acts as a bridge between the choice preference scores and realties of mobility options and requirements in different environments. Furthermore, the demonstrated method of the integration of behavioral factors, such as choice preference metrics, and mobility data acts as a strong proof of concept that these decision metrics can be used to examine the interaction between behavior and mobility trends during other public emergencies such as climate-related natural disasters. This type of finding, therefore, encourages work to understand what behavioral levers might be most useful when creating effective evacuation orders or shelter-in-place mandates. Understanding human mobility patterns during environmental emergencies such as hurricanes, floodings and wildfires is valuable for comprehending how individuals react to disasters. Understanding how decision-making processes operate under these extremes of uncertainty only enhances that knowledge. Lastly, this study shows that decision agency and choice preferences can be integrated into dynamic models of disease transmission, allowing for the prediction of new infections based on anticipated human mobility. These infection forecasts can, in turn, inform updates to mobility patterns through a feedback loop that takes evolving decision preferences into account.

## Materials and methods

### Survey data

We quantified the choice preferences and decision agency of residents of New York City through an online survey implemented using the Qualtrics platform. Details of the instrument are outlined below.

*Temporal discounting* was measured using a dynamic series of financial decisions where the amount, timeline for receipt, and difference between options were varied based on the participants previous choices - as based on previous work (Ruggeri et al., 2022). The scoring was based on a combination of responses to three baselines regarding either receiving or paying $500, and then receiving $5000 as well as dynamic follow up questions to each of those baselines. An example baseline item is “Which would you prefer? Receiving $500 right now. Receiving $550 in 12 months.” If the participant selected to take the $500 right now, the next question would increase the 12month amount to $600 to gauge the value of the delay. If the participant decided to wait for the $550, the question was asked again with the new value for the delayed option reduced to $510. These kinds of trade-offs were analyzed also for the paying $500 baseline (a loss rather than a gain), and the $5000 baseline which increased the magnitude of the initial amount. These dynamic decisions were combined estimate the temporal discounting score of the individual using the “tipping points” between the shorter and longer terms options of different magnitudes.

*Loss aversion* was measured based on the well-established method under prospect theory (Abdellaoui et al., 2007). Our measurement used 6 items that asked for preferences between different dollar amount options, each with distinct levels of certainty, framed either as a loss or a gain. For example, one item was “Which would you prefer? An 80% chance of winning $6000. A 100% chance of winning $4500.” This question was then repeated with a loss framing. The incongruency between the choices based on framing as gains or losses were used to calculate the loss aversion of each survey participant.

*Agency* was measured using a newly constructed scale that tried to understand the extent to which the individuals had agency to engage with public safety measures during the 2020 lockdowns. An example statement is “During the pandemic in 2020, the subway and/or the bus were transportation options that were…” The participants then responded to this prompt in relation to two metrics “…available close to me.” and “… realistic for me to use when I needed to.” For each metric, objective availability and subjective accessibility, the participants rated their agreement from “strongly disagree,” “disagree,” “do not know,” “agree” to “strongly agree.” There were seven main statements that formed the agency measure, meaning that all participants responded to at least 14 items. However, based on an initial statement regarding whether they worked or attended education in-person, remotely, or in hybrid format, up to two further statements could be asked of participants leading to a maximum number of 18 items. The majority answered 16 items and thus the scores were rescaled to give a unified scoring system by dividing the number of questions actual answered and multiplying by 16. The items were scored from –2 to 2 giving a total range of – 32 to 32 for the full-scale scoring.

As this was a new measure the psychometrics of the scale were checked in accordance with standard cut-off values of reliability and validity of the items (Greiff & Allen, 2018). This is not to try to establish the scale as a generalizable instrument, as it is highly context and temporally specific, but to check that the items are internally consistent enough to be scored collectively. Theoretically the scale lends itself to a unified one-factor model for analysis. This one factor model was internally consistent with both Cronbach’s α and *ω* reliability indicators being above >0.85 (Crutzen & Peters, 2017). However, while the SRMR was acceptable (0.075), the RMSEA was unacceptable (0.14), as were the overall model fit indicators (CFI = 0.73 and TLI = 0.68).

Based on the homogeneity of factor loadings (all between 0.51-0.68), and the strong theoretical motivation for a one-factor model, the modification indices for the measure were assessed. Several modification indices were high around certain items suggesting that allowing for the covariance between the availability and accessibility items within each of these statements would improve the model fit considerably. This was done in a stepwise manner starting from the covariance that had the highest modification index – the availability and accessibility of doctors/healthcare providers. Subsequently 4 other covariances were added in a stepwise manner (grocery stores, pharmacy stores, online shopping, and public transportation). At each point the psychometric properties of the scale were reassessed. After these 5 modifications the 1 factor model was estimated as being reliable (α and *ω* > 0.85), the SRMR was excellent (0.05) (the RMSEA was questionable (0.09)), and the CFI was acceptable (0.89) (the TLI was questionable (0.87)). The modified covariances had factor loadings between 0.26-0.34. These modifications appeared to improve the model fit extensively.

Only two statements did not have their item covariances stipulated in the model. These were based on the use of food delivery services (such as UberEats or InstCart) or ride-sharing apps (like Uber or Lyft). The modification indices for those item covariances were lower than other potential modifications that could have been adopted. While theoretically including them may have made for a more holistic modified model, an exploratory change to the model to do so did not improve the model fit substantively. Furthermore, it does seem that the availability and accessibility of these two more “luxury” options diverged in ways that the others did not – while these options were available to everyone, they were not realistic or accessible to many. Thus, while a one factor model is a good fit to the data as a whole, the ability of the scale to pull apart the availability and accessibility aspects of the agency of our participants during the 2020 lockdowns remains a strength.

*Demographic* variables such as gender, age, and education as well as self-reported adherence to behavioral policy changes (such as wearing masks in public or indoor socializing) during the lockdown in New York City were solicited at the end of the survey after all behavioral measures were completed.

### Data collection

In total we collected N = 2019 participants using a survey tool built on Qualtrics that was distributed by data collection company Cloud to collect a sample that spanned across the zip codes and boroughs of New York City. This was collected in a panel format and participants were compensated according to the Cloud compensation system. Additional participants were collected via snowball sampling in hard-to-reach zip codes to try and increase the spread of sampling across zip codes and boroughs. Participants had to be long term residents of their neighborhood such that they were living in the same zip code at the time of data collection (2023) as they were during the 2020 lockdown. This allows for the behavioral scores to be mapped at a sub-population level to the mobility data from that same time period. These snowballed participants were compensated with a $10 gift card for their time. The final sample of valid responses, after exclusions were applied, was N = 1344. For the purposes of our main analyses, we focused on those data that were linked to zip codes where we had a minimum of N = 15 participants and excluded other zip-codes. This left 17 MODZCTAs for our main analyses representing a total of N = 1049 participants.

All participants viewed an information sheet and consented to participation prior to completing the survey. All identifiable information, excluding the zip code itself, was removed from the data prior to analysis.

### Mobility data

The mobility data used in this study are from Advan, an open-source tracking of people’s foot traffic data that is recorded as census block level weekly patterns. We used visiting patterns as a proxy for people’s engagement with diverse activities over the course of the pandemic. Using the North American Industry Classification System (NAICS) code (20), we classified all places of interests (POI) into seven categories: i. grocery stores and pharmacies, ii. general retail, iii. arts and entertainment venues, iv. restaurants and bars, v. educational settings, vi. healthcare facilities from this dataset and vii. Others (Table S1). Our paper focuses on the categories apart from the vii. others as this category pertains to various POIs and its nature is hard to classify. Similar mobility data derived from mobile devices have been used to inform epidemic modeling (Pei et al., 2021) and quantify mobility during COVID-19 and other natural disasters (Hajlasz & Pei, 2024; Hong et al., 2021; Lu et al., 2012).

Based on these visits to POIs and classifications, we constructed a daily mobility matrix 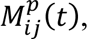 describing the number of visits originating from zip-code area *j* to POI category *p* in zip-code area *i* at day *t*. The final size of the mobility matrix for each day, across all categories, is 177 × 177, representing all zip code areas in New York City. Then we focused on the temporal “travelling out” patterns of each MODZCTA. To account for population variance, we scaled the visits with population, calculating as 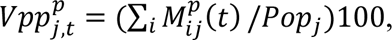 where *Pop*_*j*_ denotes the population of MODZCTA j.

The mobility data are recorded at the census tract level and in a weekly pattern. This means the crosswalk between census tract, the MODZCTA, and how we distributed the visitors across the days of the week and the known home locations is not unique. This mapping is described below in the Data analysis section). There are two other ways one can construct similar mobility matrices with these data. We tested our models on these alternative mobility matrices, see the SI, for robustness analysis.

### Data analysis and statistical analysis

The following data processing and analyses were conducted with Python languages and R software.

### Mapping between TRACT, ZIP Code, ZCTA, and MODZCTA

The data sources for this study intersected across distinct units of geographic area. The mobility and social economic data are recorded at the census tract level. Our survey data is collected at the zip code level. The COVID-19 transmission data is reported at the MODZCTA and Borough level. These data units needed to be aligned prior to analysis.

Census tracts represent small administrative areas. In contrast, ZIP Code Tabulation Areas (ZCTAs) are larger administrative units that encompass multiple tracts but are not unique to them. In most cases, a ZCTA is the same as its associated ZIP Code. However, ZCTAs and zip codes are not always the same. As not all ZCTAs have residents, the New York City Department of Health & Mental Hygiene (DOHMH) utilizes Modified ZIP Code Tabulation Areas (MODZCTAs) for monitoring COVID-19 transmission data, and certain ZCTAs that are without residents are merged in this context. To examine the crosswalk between these classifications, ZCTA to MODZCTA, in detail please review https://github.com/nychealth/coronavirus-data/tree/master/Geography-resources.

For our project, in order to make valid geo-spatial comparisons, we used the cross walk between the zip code and the TRACT provided by the Office of Policy Development and Research (https://www.huduser.gov/portal/datasets/usps_crosswalk.html). This cross walk aggregates the mobility data from the tract level to the zip code level by assigning each tract’s mobility data to the zip code area where most of its population resides. The crosswalks between zip codes and ZCTAs are then found in the GitHub repo (https://github.com/censusreporter/acs-aggregate/tree/master) from the Knight Lab. The social economic data at the ZCTA level was sourced by the R package ‘tidycensus’ (https://walker-data.com/tidycensus/index.html). Once all the data was aligned to the ZCTA level, we followed the DOHMH’s cross walk to map the data from ZCTAs to MODZCTAs.

### Generalized additive model

The generalized additive model (GAM) is a flexible nonlinear regression model, an extension of the additive model where the linear predictor is replaced by a sum of smooth functions of the predictor variables (Hastie & Tibshirani, 1986, 1987). In our analysis, we used the log-transformed weekly number of visits to different place categories as the response variable because the visit numbers spanned several orders of magnitudes. We found that the response variable quantifying weekly number of scaled visits to a given place category had strong temporal autocorrelations. The GAM model can better capture such temporal autocorrelation in the response variable by introducing nonlinear spline terms. To further alleviate the temporal autocorrelation, we further included the mobility in the prior week to the model covariates.

In this model, we allowed the covariates of mobility the week prior and the log borough case count to vary non-linearly. This allowed us to capture their possible dynamic temporal characteristics relating to established effects such as pandemic fatigue. All other covariates were estimated as linear effects as there was no theoretical foundation for their non-linearity nor did our sample have the power to effectively estimate such an unconstrained model.

Specifically, our model is constructed as follows:

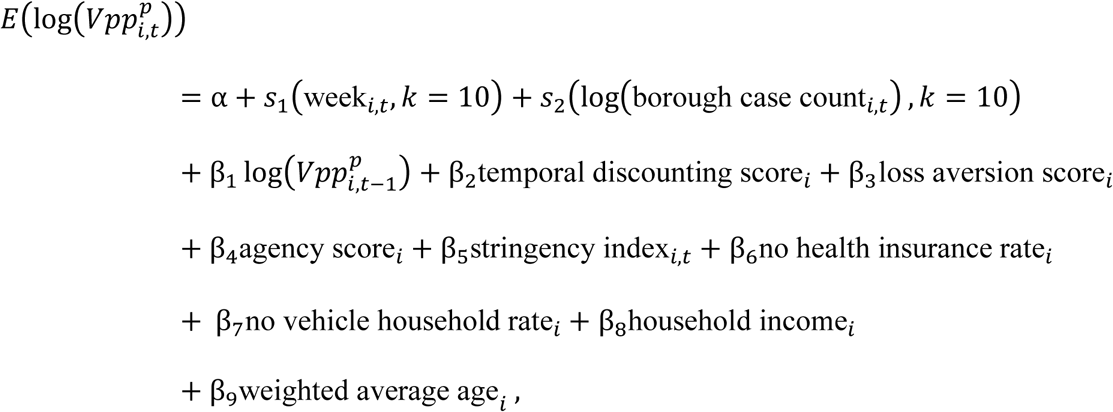

where *a* is the interception term and *s*s are the splines for the nonlinear terms, bs are the coefficients of linear terms, and *k* quantifies the flexibility of the splines. A higher value of *k* allows for a more flexible fitting of the nonlinear splines. The inclusion of 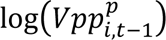 in the covariates accounts for the time dependence of the number of visits in the previous week.

We fit the model to mobility data in 17 zip-codes from 2020-03-16 to 2021-02-28 using the ‘mgcv’ R package (Wood, 2017). The R-squared values indicate the model fitting is satisfactory. We also confirmed that there was no temporal autocorrelation in the residuals for most locations. Details on the model validation and residual analysis can be found in the SI.

### Random Forest model and SHapley Additive exPlanations Importance

A Random Forest (RF) is a machine learning model that builds a “forest” of multiple decision trees and combines their results to make predictions (Breiman, 2001). The model is applicable to both classification and regression problems. In our study, we applied a RF regressor to predict the 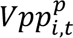 at time *t*, using three time-varying variables at time *t* − 1: week_*i*,*t*_, log(borough case count_*i*,*t*−1_), stringency index_*i*,*t*−1_, and three score variables: temporal discounting score_*i*_, loss aversion score_*i*_ and agency score_*i*_.

The initial training was performed based on the first 30 weeks (*t* ∈ [ 1, . . . , 30]) and the first prediction was made for *t* = 31. We then applied an expanding training window approach to predict the following 19 weeks, training 19 different models for each category. R-squared values are calculated using the 19 predictions across 17 MODZCTA for each category.

SHapley Additive exPlanations (SHAP) is a unified framework to explain the output of model predictions (Lundberg, 2017) by calculating SHAP values, which represent each feature’s contribution to the prediction. These values are extended Shapley values in game theory and are computed as the weighted sum of impact of a feature to the total model prediction (Shapley, 1953). The Shapley value of feature :

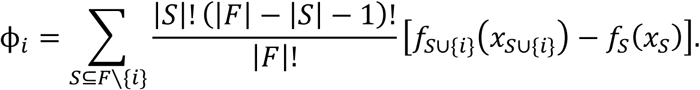

Where *S* denotes all subsets of the set *F* of all features and *f* is the prediction model.

For sensitivity analysis, we performed the prediction with adding the socio-economic variables -- no health insurance rate_*i*_,no vehicle household rate_*i*_, household income_*i*_ and weighted average age_*i*_. The performance was similar to the models with fewer variables, see the SI for the detailed results.

We used the “scikit-learn” package (Pedregosa et al., 2011) for training and prediction and “shap” package for feature importance calculation (https://github.com/shap/shap).

## Acknowledgments section

We thank Advan for sharing the foot traffic data.

## Declaration of conflicting interest

The author(s) declared no potential conflicts of interest with respect to the research, authorship, and/or publication of this article.

## Funding statement

This study was supported by funding from National Science Foundation (DMS-2229605), Centers for Disease Control and Prevention (U01CK000592 and 75D30122C14289), and Council of State and Territorial Epidemiologists (NU38OT00297).

## Ethical approval

The survey data was collected in line with the local IRB at Columbia University (Protocol: AAAU2984(M03Y01) – approved 02/28/2024). All research was performed in accordance with the guidelines set out in the Declaration of Helsinki. The IRB gave this study an exempt research status.

## Informed consent

All participants viewed an information sheet and consented in writing to participate immediately prior to completing the survey on the online platform Qualtrics. They consented to their own participation in the study; no proxy consent was obtained. The consent statement included that it related to their participation, data use, consent for their data to be used for academic research purposes, the anonymity of their data was assured, and that there were no risks to their participation. Participants were informed that the research was conducted to understand behavior during the pandemic. No vulnerable individuals were recruited for this study.

## Data availability statement

The data that support the research, including the anonymous survey data, cleaned mobility pattern and the scores, are available in GitHub repository (https://github.com/Qing1011/behaviours_epidemics). Python and R codes for this research are available in our GitHub repository (https://github.com/Qing1011/behaviours_epidemics).

## Notes

### Competing Interest Statement

The authors have declared no competing interest.

### Author Declarations

IRB of Columbia University waived ethical approval for this work and gave this study an exempt research status. (Protocol: AAAU2984(M03Y01) approved 02/28/2024)

