## Supplemental Information for "Person, place, or preference? Combining subjective and objective decision agency with choice preferences predicts behavior during major shifts in human mobility"

### A. Mobility data

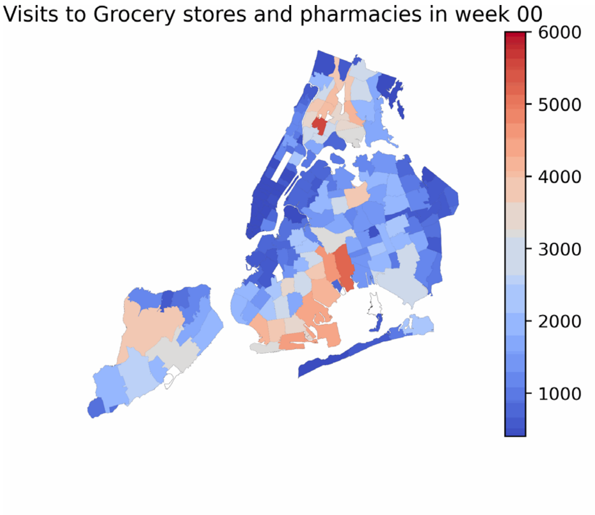


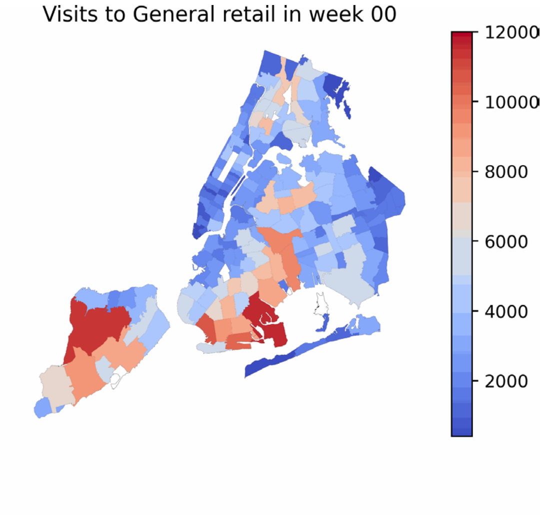


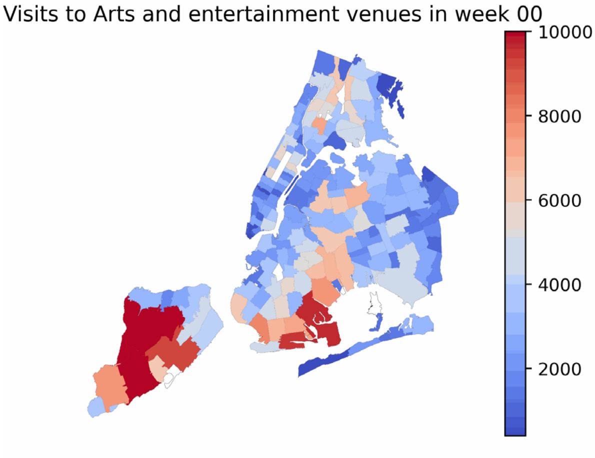


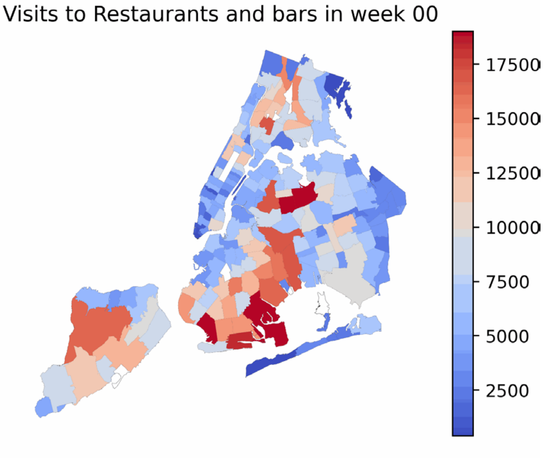


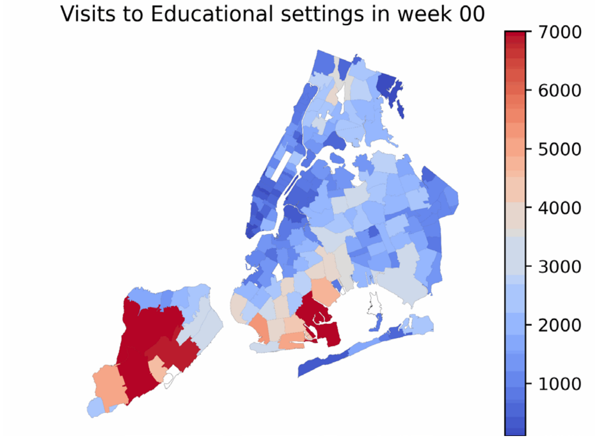


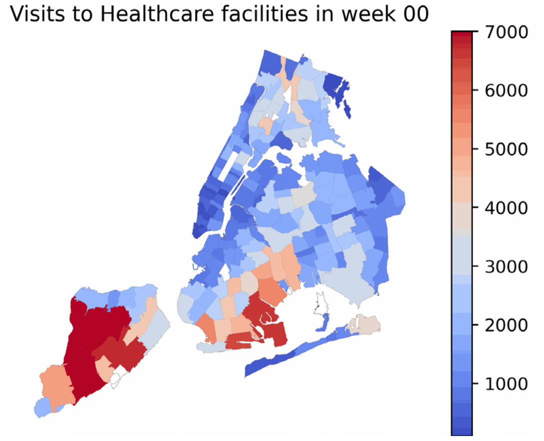

*Note.* **Movie 1 (a)** The raw visits the people reside in the colored MODZCTA paid to different types of POI from February 24, 2020 (Monday) to February 28, 2021 (Sunday) for 56 weeks.

### B. Individual temporal patterns in indoor activity choices over five periods

We asked participants about their indoor activity choices across five distinct periods during the COVID-19 pandemic:

(1) March 2020 (the start of the pandemic, before guidelines were clear, NYC cases rising rapidly).

(2) Spring 2020 (during the full lockdown, before reopening, NYC rates dropping but high number of cases still).

(3) Summer 2020 (first phase of reopening, mostly outdoor activities, NYC cases drop and remain low).

(4) Fall 2020 (indoor dining reopens, NYC cases rise slightly but overall low).

(5) Winter 2020-21 (NYC cases rise heavily, indoor dining stopped).

Participants were asked to rank their frequency of use based on the following categories, and we scored the choices from most liberal 6 to most conservative 1:

- 5: The same as before the pandemic
- 4: Multiple times each week
- 3: Weekly
- 2: Once or twice a month
- 1: Rarely, only when unavoidable

- 0: Never

Each participant provided a vector of 5 choices corresponding to their behaviors across the five periods. Using K-means clustering, we classified participants' choice patterns into five distinct classes. The following figure illustrates these patterns:

**Figure S1**

*The Indoor Activity Cluster Heatmap***.**

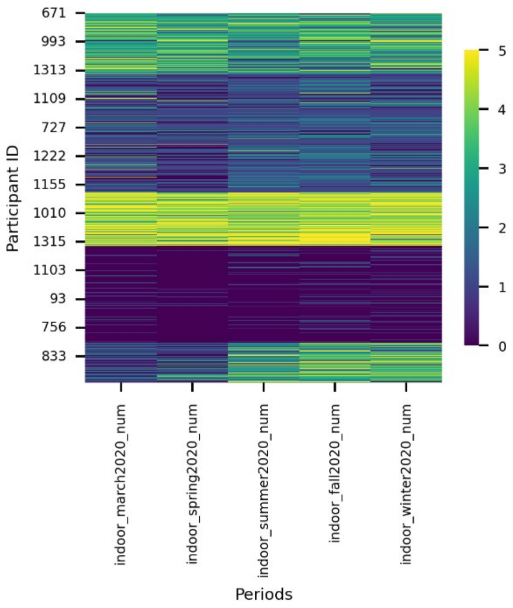


Almost one third of participants rarely or never engaged in indoor activities across all periods. About one-fifth of participants were initially conservative but increased their indoor activity frequency over time. The remaining participants consistently maintained either a high or moderate frequency of indoor activities throughout the periods. These individual-level patterns help explain the population-level observations: a decrease in indoor activities reduced the need to travel to different place categories, leading to an overall reduction in mobility. A mild increase in indoor activities after reopening contributed to a recovery in mobility.

### C. The differences in agency

**Figure S2**

*The plot for agency difference against the agency*

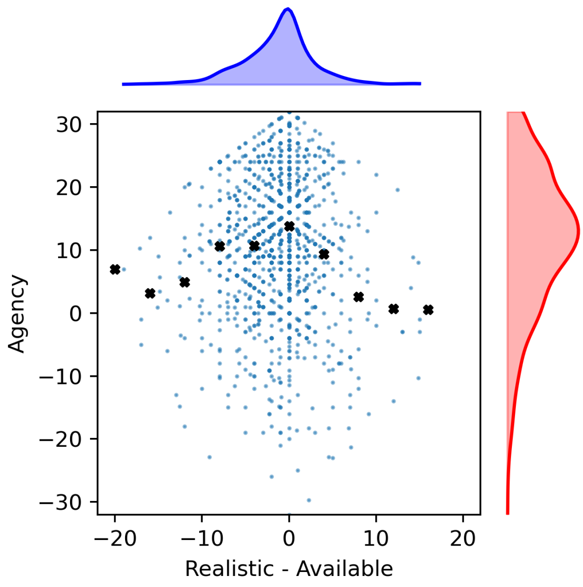


*Note.* The small blue dots are the individual participants, and the black crosses are the average value of the agency score of the bin. The bin size is 4.

The distribution of differences is skewed to the left, indicating that more people experience a situation where R<A, meaning that some services exist but are hardly affordable. Similarly, the distribution of the agency score is also skewed to the left, suggesting that, on average, more people agree or strongly agree with statements such as 'xx is available/realistic to me.' When comparing the agency scores for the same level of difference between the positive and negative sides, the positive side (R>A)—representing 'I am willing to pay something, but the services are not accessible'—is associated with lower agency scores compared to the negative side (R<A). This implies that physical or objective constraints tend to limit agency more significantly.

**Figure S3**

*Geographical distribution of subjective and objective agency for different services*

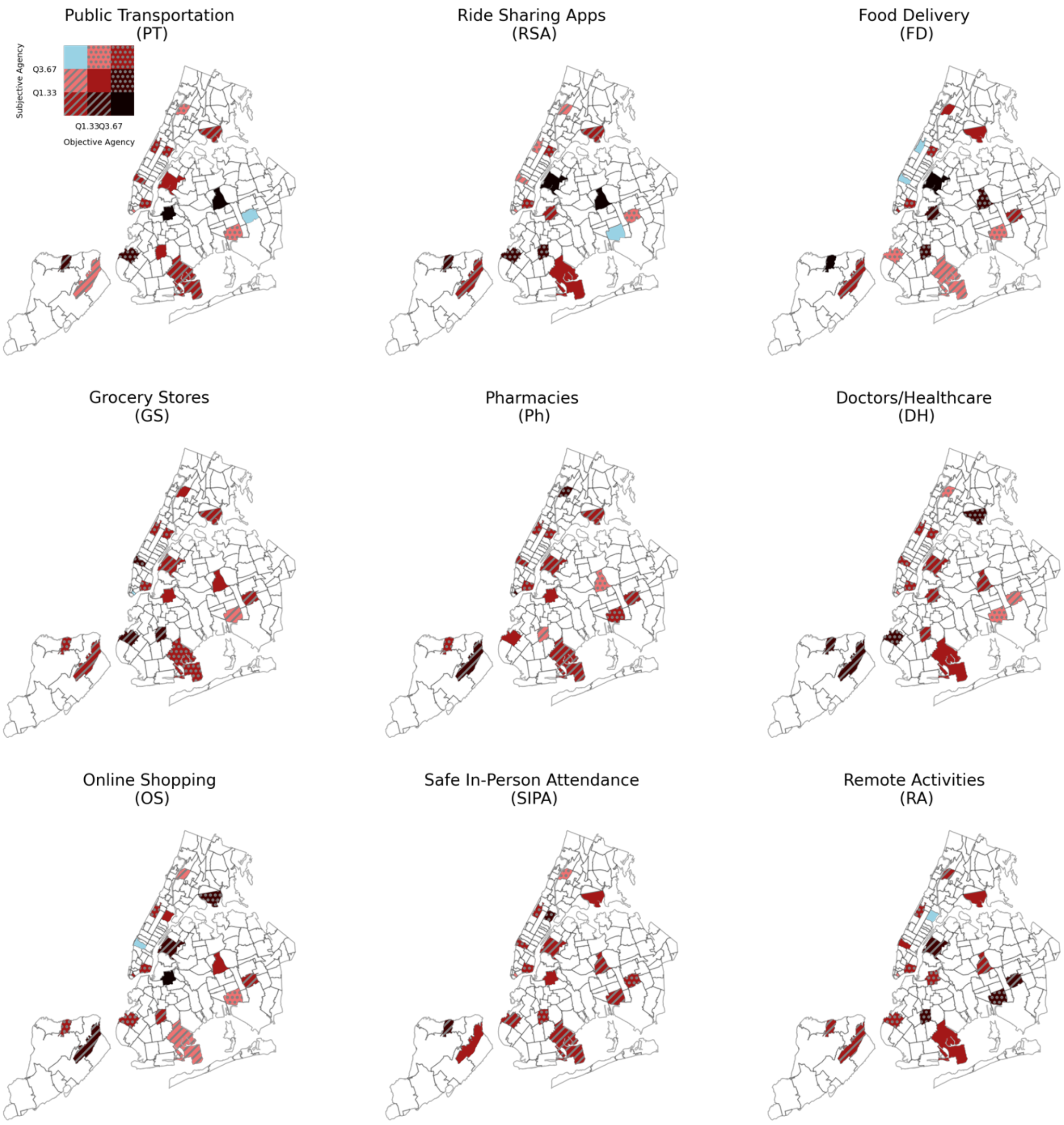


*Note.* Bivariate color scheme of mean objective and subjective agency of each service in the ZIP codes where at least 15 participants provided responses. We used quantiles to categorize the different level of objective and subjective agency scores.

### D. Data summary

#### Table S1

*Data Summary*

| Name | Description | Mean | Std | Min | P25 | Median | P75 | Max | Region | Temporal | Source | Comments |
|---|---|---|---|---|---|---|---|---|---|---|---|---|
| participants' temporal discounting score | scores calcuated for each participant based on their answers to survey questions and then we rescaled to [0-100] | 65.467 | 20.207 | 0.000 | 50.000 | 68.750 | 81.250 | 100.000 | individual | no | our survey data | before rescaling, the td score ranges from 0 to 16 |
| participants' loss aversion score | | 61.049 | 22.503 | 0.000 | 50.000 | 66.667 | 83.333 | 100.000 | individual | no | our survey data | before rescaling, the td score ranges from 0 to 6 |
| participants' agency score | | 68.019 | 17.710 | 0.000 | 57.645 | 69.444 | 80.357 | 100.000 | individual | no | our survey data | before rescaling, the td score ranges from -32 to 32 |
| Grocery and Pharmacies | weekly visits per 100 population flow out from a mod for a given type | 1.601 | 0.857 | 0.010 | 0.941 | 1.408 | 2.392 | 3.794 | MODZCTA | yes | Advan | the week starts from Monday |
| General Retail | | 3.588 | 1.975 | 0.174 | 2.150 | 2.955 | 4.818 | 9.452 | MODZCTA | yes | Advan | |
| Arts and Entertainment | | 3.513 | 1.420 | 0.572 | 2.569 | 3.224 | 4.174 | 8.726 | MODZCTA | yes | Advan | |
| Restaurants and Bars | | 6.276 | 2.245 | 1.128 | 4.600 | 5.911 | 7.670 | 12.200 | MODZCTA | yes | Advan | |
| Education | | 0.636 | 0.337 | 0.010 | 0.409 | 0.577 | 0.777 | 2.344 | MODZCTA | yes | Advan | |
| Healthcare | | 1.615 | 0.981 | 0.010 | 0.837 | 1.459 | 2.115 | 5.095 | MODZCTA | yes | Advan | |
| temporal_discounting_score | The mode of kernel density of temporal discounting scores in one MODZCTA, ranging from 0 to 100 | 66.091 | 3.386 | 58.333 | 65.278 | 67.298 | 68.750 | 69.697 | MODZCTA | no | our survey data | |
| loss_aversion_score | The mode of kernel density of loss and gain aversion scores of one MODZCTA, ranging from 0 to 100 | 57.378 | 7.221 | 50.337 | 52.020 | 54.545 | 59.596 | 71.717 | MODZCTA | no | our survey data | |
| agency_score | The mode of kernel density of agency scores of one MODZCTA, ranging from 0 to 100 | 69.247 | 5.852 | 56.897 | 66.947 | 68.939 | 70.712 | 86.111 | MODZCTA | no | our survey data | |
| week | The order of the week current week from the 16th March | 24.500 | 14.439 | 0.000 | 12.000 | 24.500 | 37.000 | 49.000 | city | yes | | |
| log_borough_case_count | Logrithsm of number of confirmed cases at borough level in one week | 6.895 | 1.827 | -2.303 | 6.140 | 6.917 | 8.139 | 9.339 | borough | yes | https://github.com/nychealth/coronavirus-data/tree/master/trends | this is from Sunday and the 6 days before March 1st 2020 |
| stringency_idx | The policy stringency index by oxford university | 71.316 | 12.511 | 14.683 | 72.220 | 72.690 | 75.000 | 82.410 | city | yes | https://www.bsg.ox.ac.uk/research/covid-19-government-response-tracker | from Monday and the 6 days after and then average |
| no_health_insurance_rate | The percent of the population who has no insurance | 0.064 | 0.031 | 0.010 | 0.038 | 0.055 | 0.089 | 0.123 | MODZCTA | no | USA census | |
| no_vehciles_householde_rate | The rate of households which do not have vehciles | 0.574 | 0.247 | 0.196 | 0.354 | 0.668 | 0.757 | 0.914 | MODZCTA | no | USA census | |
| household_income | The median value of the household income | 76826.8 | 39057.3 | 33901.0 | 49013.0 | 68801.0 | 96321.0 | 197188.0 | MODZCTA | no | USA census | |
| weighted_average_age | Averaged from the (midpoint ages * population of the range) / total populations | 38.156 | 3.553 | 31.819 | 36.089 | 38.904 | 40.039 | 44.374 | MODZCTA | no | USA census | |

### E. Correlations between individual behavioral scores and other factors

**Table S2**

| | **Temporal Discounting** | **Loss Aversion** | **Agency** |
|---|---|---|---|
| Age | -0.025 | -0.006 | **0.207** |
| Education | **-0.054** | **-0.104** | **0.196** |
| Employment | -0.042 | **-0.074** | **0.135** |
| Financial 2020 | **-0.102** | 0.013 | **0.193** |
| Financial now | **-0.191** | -0.034 | **0.165** |
| Financial change | **-0.096** | **-0.065** | **0.194** |
| Credit and Debt | **-0.149** | 0.019 | **0.092** |
| Expectation | 0.043 | 0.011 | **0.223** |

*Note.* Pearson correlation table between scores and personal information (N = 1341, there are 3 participants did not provide full personal information). Correlation coefficients are shown in bold if the *p*-value is less than 0.05, indicating statistical significance.

### F. Full regression results

**Figure S4**

*Partial dependence plot for all the variables in the model*

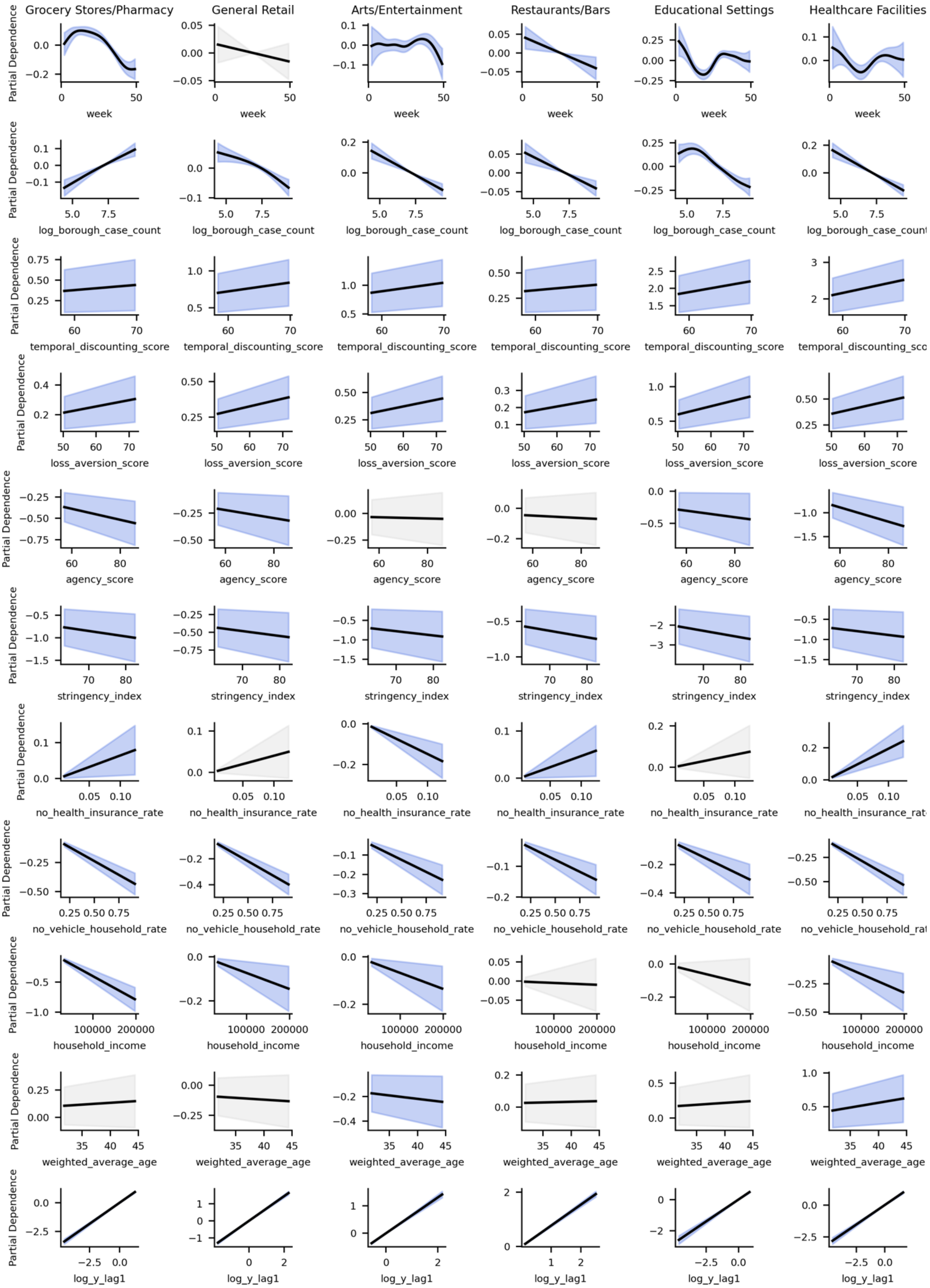
Grocery Stores/Pharmacy
General Retail
Arts/Entertainment
Restaurants/Bars
Educational Settings
Healthcare Facilities
Partial Dependence
week
log_borough_case_count
temporal_discounting_score
loss_aversion_score
agency_score
stringency_index
no_health_insurance_rate
no_vehicle_household_rate
household_income
weighted_average_age
log_y_lag1

*Note.* Each column represents the model results for a different category, while each row corresponds to a specific variable. Variables that are statistically non-significant in the GAM are in light gray shades.

### G. GAM Model validation

**a) residual analysis**

Different POI categories can have different estimated degree of freedoms (EDFs) for the two variables. We chose k = 10 for both variables to avoid the overfitting in the main results and performed the residual checks, see the results here:

**Figure S5**

*Plots for GAM model check for different place categories*

Grocery_and_Pharmacy

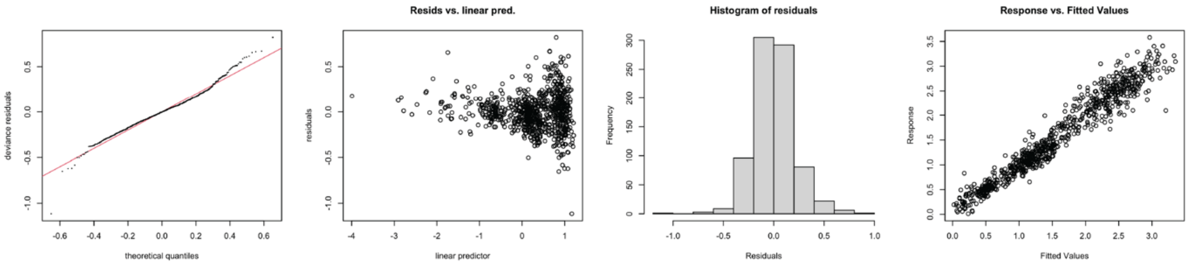


General_Retail

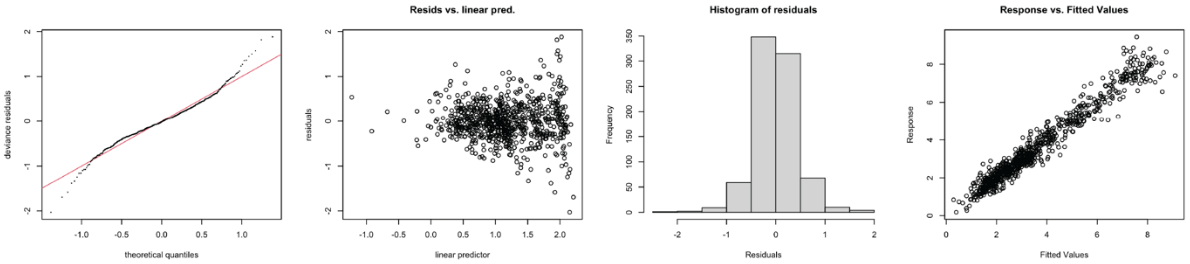


Art_and_Entertainment

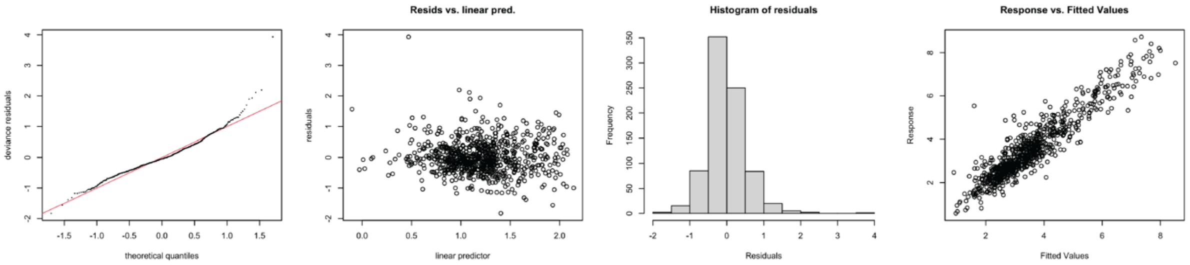


Restaurant_and_Bar

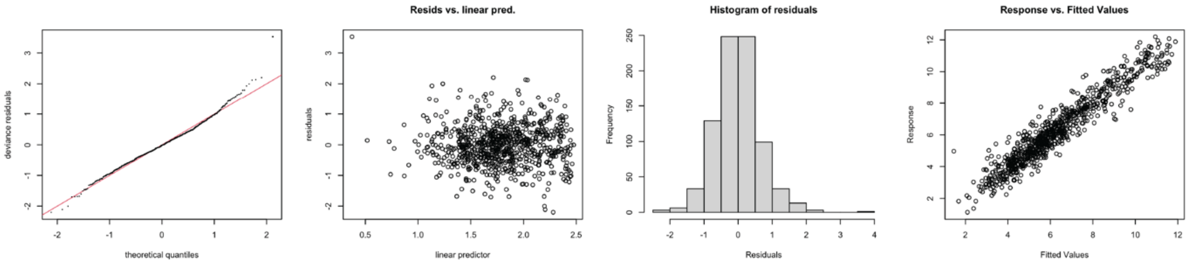


Education

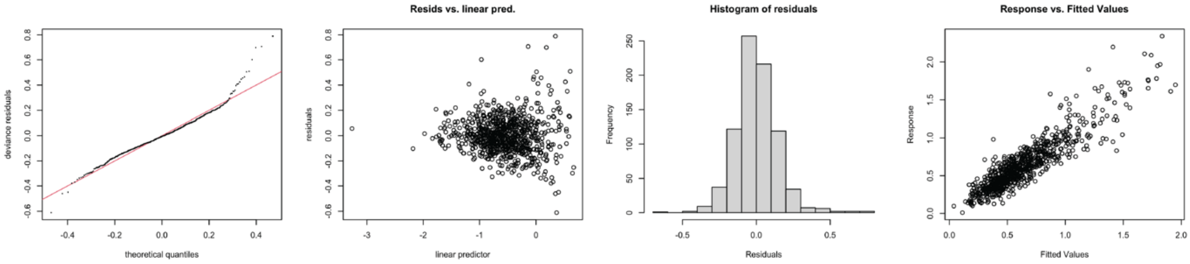


Healthcare

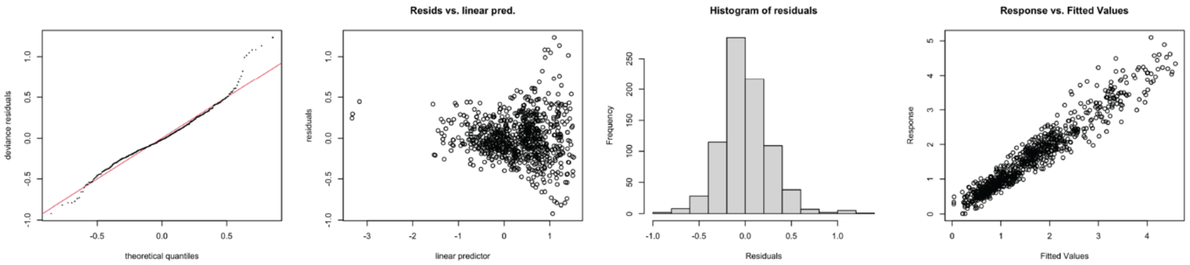

**b) Temporal correlation analysis**

We further grouped the residuals within zip codes and checked the temporal autocorrelations within each zip code using Durbin-Watson test. The results are shown in the following. Results indicate that no significant temporal correlations exist in the residuals in all zip codes.

**Figure S6**

*Plots for auto correlation check of GAM model in each zip code*

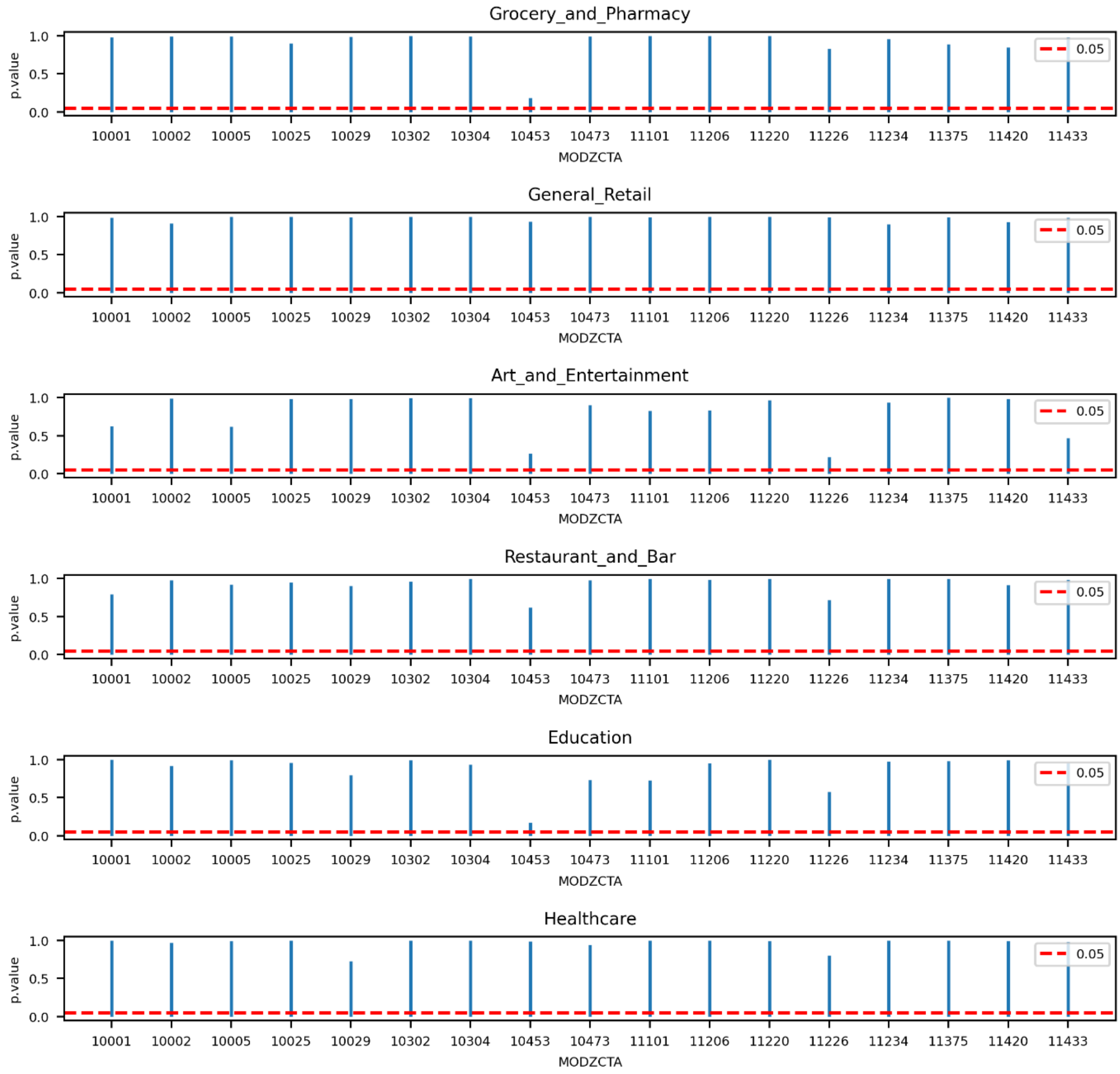


**c) Sensitivity analysis**

We additionally performed the models on k = 9 and k = 11, the splines are similar:

**Figure S7**

*Plots for partial dependences of nonlinear terms when k = 9*

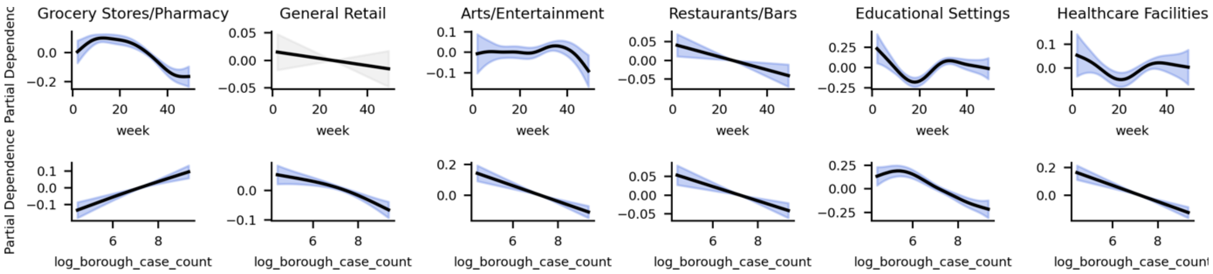


**Figure S8**

*Plots for partial dependences of nonlinear terms when k = 11*

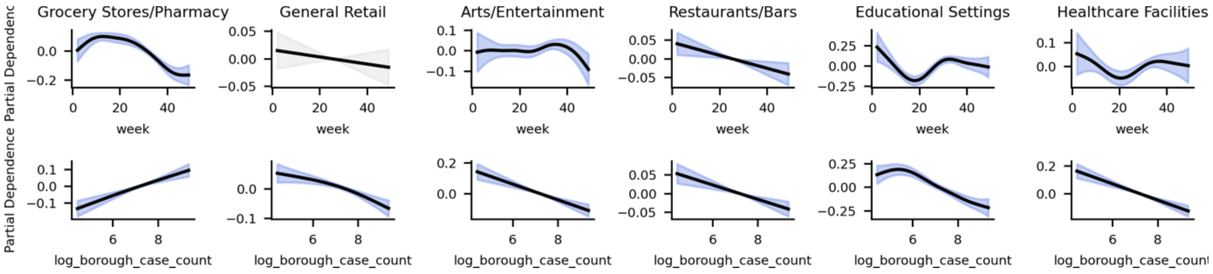


**c) robustness analysis**

The mobility data is aggregated from the census tract level to the MODZCTA level. We map each census tract to the MODZCTA it belongs to, or to the one where it has the highest residence ratio. Additionally, when assigning visitors to their home census tracts, we excluded those whose home locations were not recorded. To account for those factors, we propose two additional methods for aggregating the data: i) Mapping Visits by Residence Ratios: Attribute visits from the census tract level to the MODZCTA level according to residence ratios; ii) Assigning Lost Visitors: Distribute visitors without recorded homes proportionally to the known home census tracts.

We test our models on the two above mobility aggregating methods, see the regression results in the following:

i) one census tract to multiple MODZCTA mapping

**Figure S9**

*Partial dependence plots for GAM when processing data with a ‘1 to more’ mapping method*

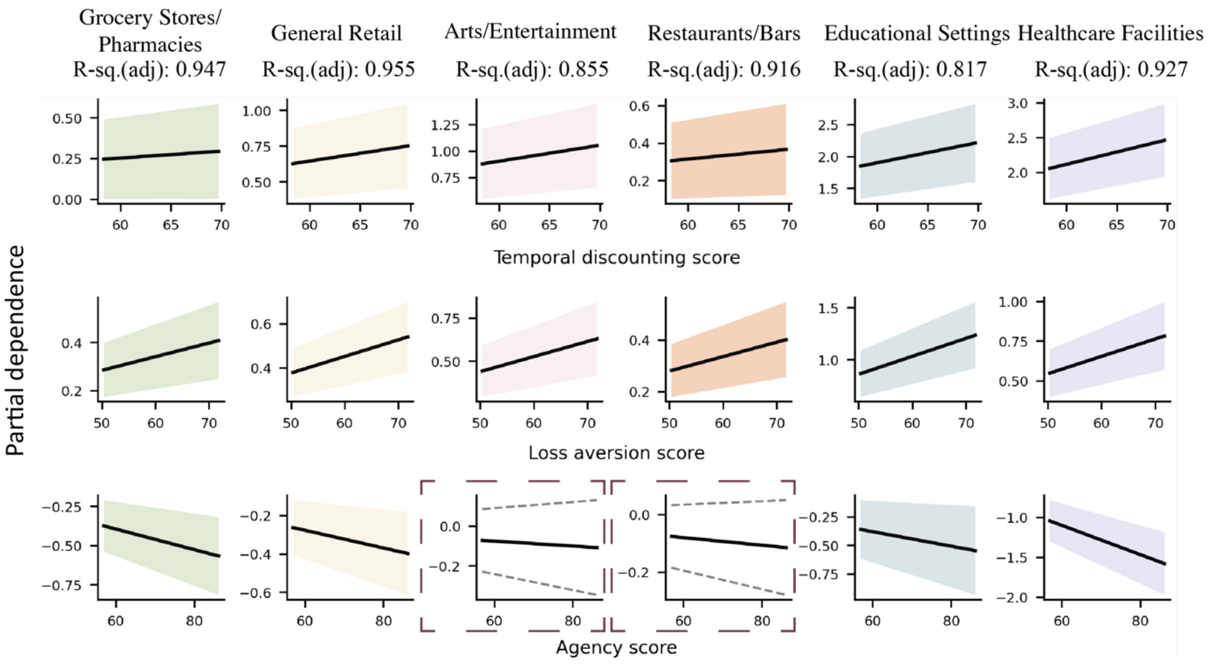


ii) assign loss visitors

**Figure S10**

*Partial dependence plots for GAM when processing data with assigning the unknown visitors according to the known ratio*

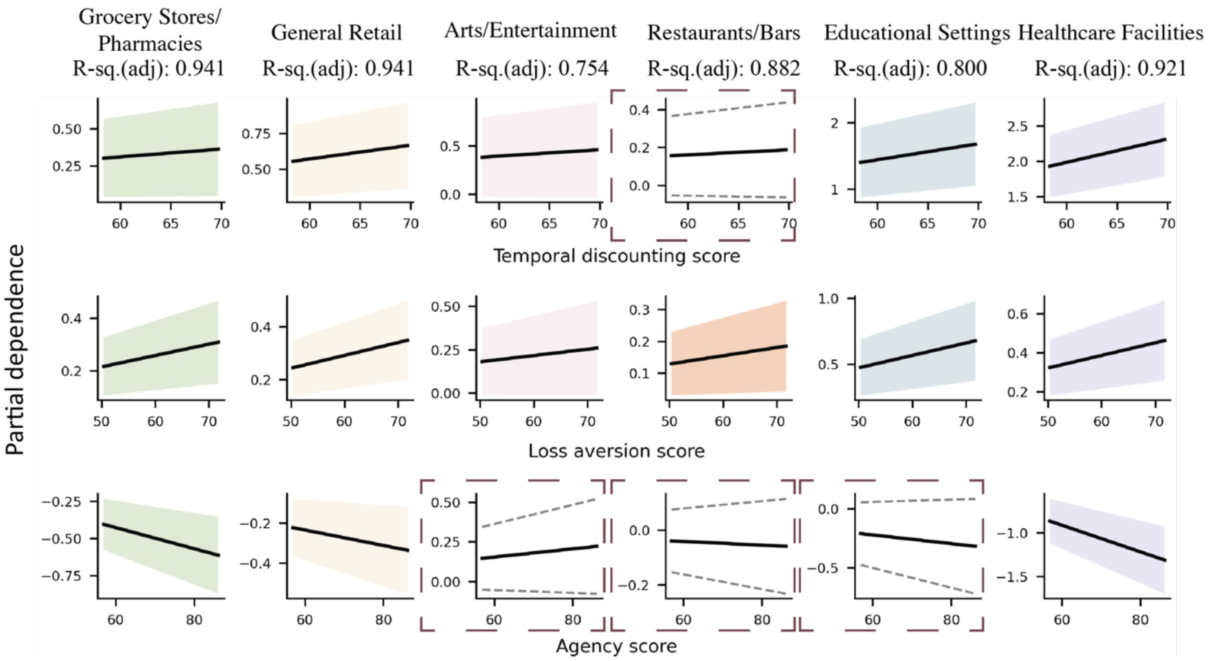


The significance of temporal discounting score for the Restaurants and bars and the agency score for Educational settings were changed. This data processing assumed that the visitors' home zip codes were distributed uniformly, or the unknown could be presented by the known part which might be too strong in our study. Thus, we chose not to use this method.

**H.** Random Forest (RF) model

a) To further evaluate the prediction results of RF models. We plotted the predicted values with the true values against the time for each category and location.

**Figure S11**

*The predictions and true values of weekly visits per 100 per capita for each category across 17 MODZCTAs*

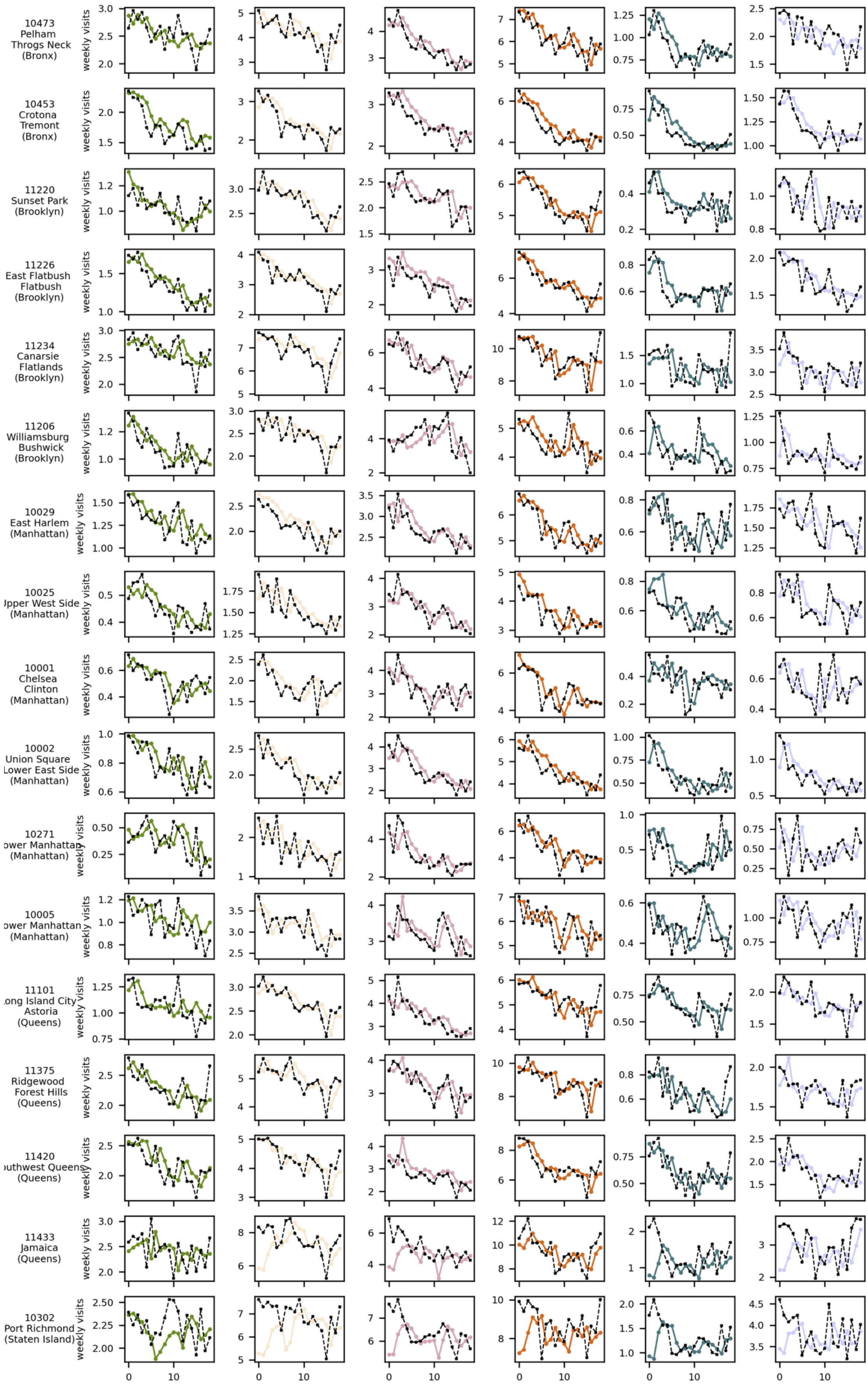
10473
Pelham
Throgs Neck
(Bronx)
10453
Crotona
Tremont
(Bronx)
11220
Sunset Park
(Brooklyn)
11226
East Flatbush
Flatbush
(Brooklyn)
11234
Canarsie
Flatlands
(Brooklyn)
11206
Williamsburg
Bushwick
(Brooklyn)
10029
East Harlem
(Manhattan)
10025
Upper West Side
(Manhattan)
10001
Chelsea
Clinton
(Manhattan)
10002
Union Square
Lower East Side
(Manhattan)
10271
Lower Manhattan
(Manhattan)
10005
Lower Manhattan
(Manhattan)
11101
Long Island City
Astoria
(Queens)
11375
Ridgewood
Forest Hills
(Queens)
11420
Southwest Queens
(Queens)
11433
Jamaica
(Queens)
10302
Port Richmond
(Staten Island)
weekly visits
0
10

*Note.* Each column corresponds to a specific category, while each row represents a location. The colored lines indicate the predicted values from the Random Forest (RF) models, and the dashed black lines represent the true value.

Most locations exhibited reasonable prediction results. However, certain locations, such as MODZCTA 11433 (Jamaica) and 10302 (Port Richmond), showed a noticeable mismatch between the predicted and true values. This suggests that the selected six features were insufficient to fully explain the variation in visits to different categories in these areas.

b) We built the RF models using the same sets of the variables as the GAM model. The prediction results are presented in the Table S3

**Table S3**

*Results of the Random Forest model for predicting weekly visits per 100 capita using ten variables*

| CATEGORIES | MEAN SQUARED ERROR (MSE) | MEAN ABSOLUTE ERROR (MAE) | R-SQUARED | MEAN ABSOLUTE *PERCENTAGE* ERROR (MAPE) |
|---|---|---|---|---|
| GROCERY STORES/PHARMACY | 0.03 | 0.14 | 0.95 | 13.20 |
| GENERAL RETAIL | 0.21 | 0.32 | 0.94 | 9.80 |
| ARTS/ENTERTAINMENT | 0.23 | 0.37 | 0.86 | 11.44 |
| RESTAURANTS/BARS | 0.44 | 0.50 | 0.89 | 8.71 |
| EDUCATIONAL SETTINGS | 0.04 | 0.13 | 0.74 | 21.79 |
| HEALTHCARE FACILITIES | 0.09 | 0.21 | 0.90 | 15.86 |

*Note.* These variables include the six presented in the main study (temporal discounting score, loss aversion score, agency score, week index, stringency index lag1, and log borough case count lag1) as well as four additional socio-economic variables: no health insurance rate, no vehicle household rate, household income, and average age.

Adding additional variables did not significantly improve the predictions, except for a noticeable increase in the R-squared value for educational settings, which rose from 0.60 to 0.74. For other categories, the R-squared values increased by approximately 0.1–0.2. This suggests that choice preferences and decision-making agencies captured similar variations as the socio-economic data in explaining weekly visits to most categories, except for educational settings. The socio-economic data may provide additional insights into mobility patterns specifically related to educational settings.
